# Decoding Dementia from Sweat: A Proof-of-Concept Label-Free Raman Fingerprinting of EVs

**DOI:** 10.64898/2026.09.25.26364001

**Authors:** Aleem Ud Din, Subodh Sharma, Minna Mannerkorpi, Elham Aryapour, Simo Saarakkala, Johanna Krüger, Nsrein Ali

## Abstract

Dementia is a major global health challenge; however, current diagnostic approaches often rely on invasive or costly procedures. Non-invasive molecular approaches using Raman spectroscopy and sweat-derived extracellular vesicles (EVs) may provide an accessible strategy for characterizing dementia-associated molecular changes. To determine whether Raman spectral profiling of sweat EVs can discriminate individuals with dementia from cognitively healthy controls (HC) and distinguish between Alzheimer disease (AD) and frontotemporal dementia (FTD) including the assessment of a sex-specific spectral signatures.

This proof-of-concept cross-sectional study included 88 participants: 26 HC, 37 patients with AD, and 25 patients FTD, comprising female and male participants. Sweat EVs were isolated from sweat patches and characterized using Raman spectroscopy. Differences in Raman spectral signatures between groups and diagnostic performance assessed by receiver operating characteristic analysis.

Distinct Raman spectral fingerprints were identified in sweat EVs from dementia patients compared with HC. Participant-level Raman classification discriminated dementia patients from HC with an AUC of 0.76 in the full cohort and 0.83 in a balanced sensitivity analysis. Distinct spectral differences were also observed between AD and FTD, achieving modest classification performance of 0.73 AUC. Sex-stratified analyses further revealed Raman spectral heterogeneity between diagnostic groups, highlighting additional molecular differences that were not observed in the overall cohort.

These findings provide proof-of-concept evidence that Raman spectroscopy of sweat EVs can identify dementia-associated molecular signatures potentially related to differences in protein, lipid, and metabolic composition. Furthermore, Raman profiling showed modest ability to differentiate AD from FTD. As a non-invasive approach, our strategy may represent a promising platform for molecular characterization of dementia, warranting validation in larger longitudinal cohorts.

## Introduction

Dementia affects approximately 57 million people worldwide and is expected to increase with population aging ^1–3^. Alzheimer’s disease (AD) is the most common cause of dementia, whereas frontotemporal dementia (FTD) is an important cause of young-onset dementia. AD and FTD can share clinical features, complicating differential diagnosis ^4–10^. Although established amyloid- and tau-based biomarkers support the diagnosis of AD, validated fluid biomarkers for FTD remain limited ^11,12^. The emergence of disease-modifying therapies for biomarker-confirmed AD further highlights the need for accessible and noninvasive approaches to molecular characterization of neurodegenerative disease ^13–16^. Sex-related biological differences may also contribute to heterogeneity in dementia and warrant consideration when evaluating molecular biomarkers ^17–23^.

Raman spectroscopy provides label-free biochemical fingerprints and, when combined with machine-learning analysis, can identify disease-associated molecular patterns ^24–27^. Extracellular vesicles (EVs) in accessible biofluids, including sweat, contain proteins, lipids, nucleic acids, and metabolites and may provide disease-related molecular information ^28–33^. However, sweat EVs remain largely unexplored in neurodegenerative diseases.

Building on our previously reported workflow for sweat EV analysis ^28,29^, we investigated whether Raman profiles of sweat EVs differed among cognitively healthy controls (HC) and participants with AD and FTD. We further evaluated whether these profiles could distinguish HC from dementia and AD from FTD and explored whether the spectral patterns differed by sex. To our knowledge, this is the first study applying Raman-based molecular profiling to sweat EVs in neurodegenerative dementia, supporting a non-invasive approach to early dementia detection and stratification.

## Methods

### Study Cohorts and Clinical Evaluation

This proof-of-concept study received ethical approval from the Northern Ostrobothnia Hospital District Ethics Committee (EETMK: 210/2001). Written informed consent was obtained from all participants who joined the study on a voluntary basis. The cohort comprised altogether 88 individuals, including newly diagnosed dementia patients aged 45–68 years at the memory outpatient clinic of the Oulu University hospital and HC, recruited between 2023 and 2026. Diagnoses were confirmed by neurologists specialized in dementia and neurodegenerative diseases according to established diagnostic criteria^34–36^. Each participant underwent a thorough clinical evaluation that included a neurological examination, structured interviews with patients and caregivers, brain magnetic resonance imaging (MRI), and comprehensive neuropsychological assessment. Cerebrospinal fluid (CSF) biomarkers for AD and/or fluorodeoxyglucose positron emission tomography (FDG-PET) imaging were performed when clinically needed. Participants in the HC group were confirmed to be cognitively healthy as indicated by normal Consortium to Establish a Registry for Alzheimer’s Disease neuropsychological battery (CERAD-NB) test results^37^, absence of subjective memory complaints, and preserved daily functioning. Of the 37 patients with early-onset Alzheimer’s disease (EOAD), the diagnosis was confirmed by CSF AD biomarkers in 34 cases. In 2 patients for whom lumbar puncture was declined or contraindicated, plasma p-tau217 was used in 1 patient and amyloid PET was performed in 1 patient. One patient was diagnosed clinically without CSF biomarkers, plasma p-tau217, or amyloid PET. The FTD group comprised 25 patients: 18 with behavioral variant frontotemporal dementia (bvFTD), 5 with primary progressive aphasia, 2 with right temporal variant FTD. *C9orf72* repeat-expansion (*C9exp*) testing was available for all 25 participants; 12 were *C9exp -*carriers, 12 non-carriers, and 1 had an intermediate repeat length of uncertain significance. Study participants were categorized into two diagnostic groups (37 AD cases and 25 FTD cases), in addition to the HC (n=26) (Table 1).

**Table 1.** Demographic and Clinical Characteristics of Study Participants. Data are means ± SD. Characterization of participant number, age, sex, time from dementia diagnosis to sample collection, and the diagnostic biomarker information were presented. HC (n=26), AD (n=37), and FTD (n=25). HC, cognitively healthy controls; AD, Alzheimer disease; FTD, frontotemporal dementia; Aβ42, amyloid-β 42; CSF, cerebrospinal fluid; FDG-PET, fluorodeoxyglucose positron emission tomography; MRI, magnetic resonance imaging; p-tau, phosphorylated tau; p-tau217, phosphorylated tau 217; SD, standard deviation.

| Variable | HC | AD | FTD |
| --- | --- | --- | --- |
| Participants, n | 26 | 37 | 25 |
| Age, years, mean ± SD | 60.0 ± 9.2 | 62.2 ± 4.4 | 61.2 ± 5.0 |
| Sex, female/male, n | 15/11 | 20/17 | 14/11 |
| Time from dementia diagnosis to sample collection, years, mean ± SD | Not applicable | Female: 0.7 ± 0.5;<br>Male: 1.0 ± 0.5 | Female: 1.01 ± 0.40;<br>Male: 0.94 ± 0.39 |
| Diagnostic biomarker information | Not applicable | CSF Aβ42, total tau, p-tau; brain MRI; plasma p-tau217 | Brain MRI, FDG-PET, and *C9orf72* repeat mutation testing |

### Extracellular vesicles isolation and characterization from sweat patches

EVs were isolated from sweat-patch samples using our recently published protocol ^28,29^. Following filtration and ultracentrifugation, the resulting pellets were resuspended in 60 µL of 1× phosphate-buffered saline (PBS) and stored at −70 °C until further analysis. EV concentration and morphology were characterized using nanoparticle tracking analysis (NTA) and electron microscopy (EM), respectively. Detailed experimental procedures are provided in the eMethods in the Supplement.

### Raman spectroscopy sample preparation and measurements parameters

Prior to measurement, 5 µL of each sweat EV sample was deposited onto an aluminium mirror substrate and allowed to air-dry for at least 45 minutes. Raman spectra was acquired using a DXR2xi Raman Imaging Microscope (Thermo Fisher Scientific) with a 785 nm excitation laser, a 50x objective, and a 50-µm slit aperture. Each acquisition used a 1-second exposure averaged over 10 scans, with laser power set between 26.5 and 27.0 mW. Approximately 160–200 spectra were collected per sample, mainly from the peripheral regions of the air-dried droplet.

### Spectral preprocessing and data analysis after Raman imaging

Raw Raman spectra underwent spectral quality filtering, cosmic-ray despiking, spectral clipping/refitting, Savitzky-Golay smoothing, Whittaker asymmetric least-squares baseline correction, peak-based normalization, and mean-background subtraction. Background spectra acquired from aluminium substrate underwent the same core preprocessing steps. The resulting background-subtracted spectra were used for downstream statistical and machine-learning analysis. A schematic overview of the preprocessing workflow, is provided in eFigure 1 in the Supplement, with additional implementation details provided in the eMethods in the Supplement.

**Figure 1.**
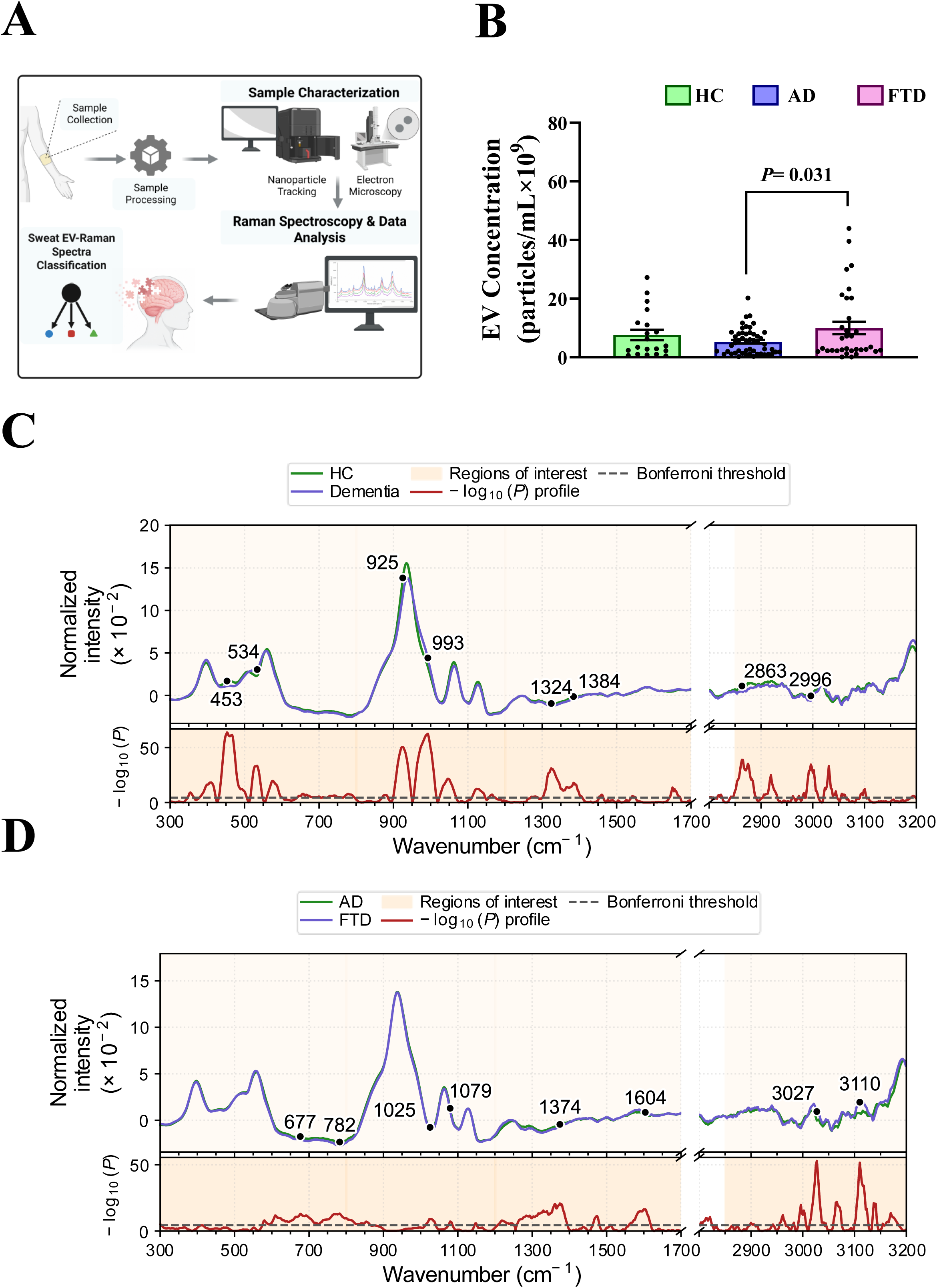
Clinical and experimental pipeline for characterizing sweat EVs and Raman spectral profiles in cognitively healthy controls, AD, and FTD. **A**, Schematic overview of the experimental workflow, including sweat EV characterization, Raman sample preparation, and spectral analysis. **B**, Quantitative analysis of EV concentration (n x 10^9^ particles/mL) in HC, AD, and FTD. Statistical differences among the groups were evaluated using ordinary one-way ANOVA followed by Tukey’s post hoc multiple comparison test. \**P*<0.05. **C**, Mean normalized Raman spectra for HC participants and participants with dementia across the fingerprint (300–1700 cm^-1^) and high-wavenumber (2800– 3200 cm^-1^) regions, with the corresponding wavenumber-wise −log₁₀(*P*) profile from 2-sided Welch independent-samples *t* tests. **D**, Corresponding Raman spectral and statistical comparison between participants with AD and FTD. In panels **C** and **D**, shaded areas indicate reporting regions, dashed horizontal lines indicate the Bonferroni-corrected significance threshold, and labeled wavenumbers denote selected significant local maxima used for subsequent analyses. EV, extracellular vesicle; HC, cognitively healthy controls; AD, Alzheimer disease; FTD, frontotemporal dementia. Graphical abstract has been created in BioRender. *Sharma, S. (2026)* https://BioRender.com/mevcci8.

### Statistical analysis of Raman spectral differences

Group-level Raman spectral differences were assessed using the background-subtracted spectra. Comparisons were performed between HC and dementia, between AD and FTD, and separately within female and male participants. At each wavenumber, normalized Raman intensities were compared between groups using a 2-sided Welch independent-samples *t* tests, within Bonferroni correction applied across the tested wavenumber points. Resulting P values were transformed to −log₁₀(*P*) to generate wavenumber-wise statistical significance profiles. Representative significant Raman bands were selected from local maxima exceeding the Bonferroni-corrected threshold with up to 2 local maxima separated by at least 25 cm^-1^ selected within each reporting region. Because multiple spectra were obtained from each participant, these analyses were considered exploratory spectrum-level analyses. Participant-level discrimination was evaluated separately using the machine-learning procedure described below. Additional statistical and visualization details are provided in the eMethods in the Supplement.

### Machine-Learning Classification of Raman Spectra

Machine-learning classification was performed using background-subtracted sweat EV Raman spectra for HC vs dementia (AD and FTD combined) and AD vs FTD, including sex-stratified analyses. Model evaluation used stratified group cross-validation with participants as the grouping variable, ensuring that spectra from the same participant were not included in both training and test sets. Up to 5 folds were used, with fewer folds when required by the number of participants in the smallest class. Within each fold, spectra underwent standard normal variate normalization, standard scaling, and principal component analysis (PCA), followed by logistic regression or a linear support vector classifier. Both classifiers used balanced class weighting, and the number of PCA components was tuned within the modelling framework. All preprocessing and model fitting steps were implemented within the cross-validation pipeline to minimize information leakage. Spectrum-level classifier scores were averaged for each participant to obtain participant-level decision scores. Performance was evaluated using area under the receiver operating characteristic curve (AUC), accuracy, balanced accuracy, sensitivity, specificity, precision, and F1 score. Models were ranked by mean cross-validated AUC, followed by balanced accuracy and accuracy. Balanced sensitivity analyses were also performed while retaining all spectra from each selected participant. For the overall HC-vs-dementia analysis, 25 HC participants and 25 participants with dementia were included, with AD and FTD representation and sex distribution matched to the HC group as closely as possible. Additional implementation and balancing details are provided in the eMethods in the Supplement.

## Results

### Sweat EV yield and their Raman fingerprints distinguish dementia and its subtypes

Following sweat-patch collection (Figure 1A), EVs were characterized. NTA showed that most EVs ranged from 100 to 500 nm, with the predominant population between 100 and 250 nm (eFigure 2A in the Supplement). Electron Microscopy confirmed the presence of EV-like structures in sweat-patch samples, with human keratinocytes (HaCaT) serving as a positive control as previously described (41). No EV-like structures were observed in negative controls (eFigure 2B in the Supplement). Sweat EV concentrations differ across diagnostic groups. Mean EV concentrations were lowest in participants with AD (n=37; 5.3 x 10^9^ particles/mL), followed by HC participants (n=26; 7.6 x 10^9^ particles/mL), and participants with FTD (n=25; 10 x 10^9^ particles/mL). EV concentration was significantly higher in the FTD group than in the AD group (*P*= 0.0317), whereas differences between FTD and HC (*P*= 0.55) and AD and HC (*P*= 0.53) were not significant (Figure 1B).

Raman spectra were subsequently acquired from sweat EV samples collected across the fingerprint (300–1700 cm^-1^) and high-wavenumber (2800–3200 cm^-1^) regions. Mean spectra and corresponding -log^10^*P* profile are shown in Figure 1C. When comparing HC with the combined dementia (AD and FTD), significant group differences were identified at 453, 534, 925, 993, 1324, 1384, 2863, and 2996 cm^-1^. At 453, 925, 1324, 1384, 2863, and 2996 cm^-1^, normalized Raman intensity was higher in HC than in dementia, whereas signals at 534 and 993 cm^-1^ signals were higher in dementia (Figure 1C). Thus, group differences extended beyond the fingerprint region into the high-wavenumber range, including C-H stretching-associated signals. Because mean spectra do not capture individual-spectrum variability, intensity distributions at the selected wavenumbers were additionally assessed using all available spectra (eFigure 3 in the Supplement), demonstrating group shifts and overlap between HC and dementia groups. The corresponding mean ± SEM values and exact *P* values are provided in eTable 1 in the Supplement. The same analysis was then performed within the dementia cohort, comparing AD and FTD. Mean spectra and corresponding −log10*P* profiles are shown in Figure 1D. Eight selected significant Raman bands were identified at 677, 782, 1025, 1079, 1374, 1604, 3027, and 3110 cm^-1^. AD showed higher intensities at 677, 782, 1025, 1079, and 1374 cm^-1^, whereas FTD showed higher intensities at 1604, 3027, and 3110 cm^-1^. The smallest P values were observed at 3027 and 3110 cm^-1^ in the high-wavenumber region (Figure 1D). Spectrum-level distributions at these bands are shown in eFigure 4 in the Supplement, with corresponding mean ± SEM and *P* values provided in eTable 2 in the Supplement.

### Sex-stratified Raman heterogeneity distinguishes dementia and its subtypes in sweat

The cohort was stratified by sex to compare sweat EV Raman profiles between HC and dementia groups. The female subgroup comprised 49 participants (15 HC, 20 AD, and 14 FTD), and the male subgroup comprised 39 participants (11 HC, 17 AD, and 11 FTD). In females, selected significant Raman bands were identified at 455, 532, 927, 973, 1255, 1567, 2863, and 3027 cm^-1^ (Figure 2A). Normalized intensities were higher in HC at 455, 927, 1255, and 2863 cm⁻¹ and higher in dementia at 532, 973, 1567, and 3027 cm⁻¹. In males, selected significant bands were identified at 655, 788, 1033, 1162, 1315, 1527, 2919, and 2998 cm^-1^ (Figure 2B). Intensities were higher in HC at all selected bands except 1527 cm^-1^, which was higher in dementia. These findings demonstrated distinct sex-stratified Raman patterns between HC and dementia. Spectrum-level intensity distributions are shown in eFigure5 in the Supplement, with corresponding mean ± SEM values and exact P values provided in eTable3 in the Supplement. We next performed sex-stratified comparisons between AD and FTD. Among 34 female participants with dementia (20 AD and 14 FTD), significant differences were observed at Raman bands 468, 547, 904, 1147, 1293, 1361, 3025, and 3110 cm^-1^ (Figure 2C). AD was associated with higher intensities at 468, 1147, 1293, and 1361 cm^-1^, whereas FTD was associated with higher intensities at 547, 904, 3025, and 3110 cm^-1^. Among 28 male participants with dementia (17 AD and 11 FTD), significant differences were observed at Raman bands 536, 775, 885, 1033, 1473, 1604, 2998, and 3029 cm^-1^ (Figure 2D). AD was associated with higher intensities at 536, 775, 885, 1033, 1473, and 2998 cm^-1^, whereas FTD was associated with higher intensities at 1604 and 3029 cm^-1^. Spectrum-level intensity distributions are shown in eFigure 6 in the Supplement, with corresponding mean ± SEM and exact *P* values provided in eTable 4 in the Supplement.

**Figure 2.**
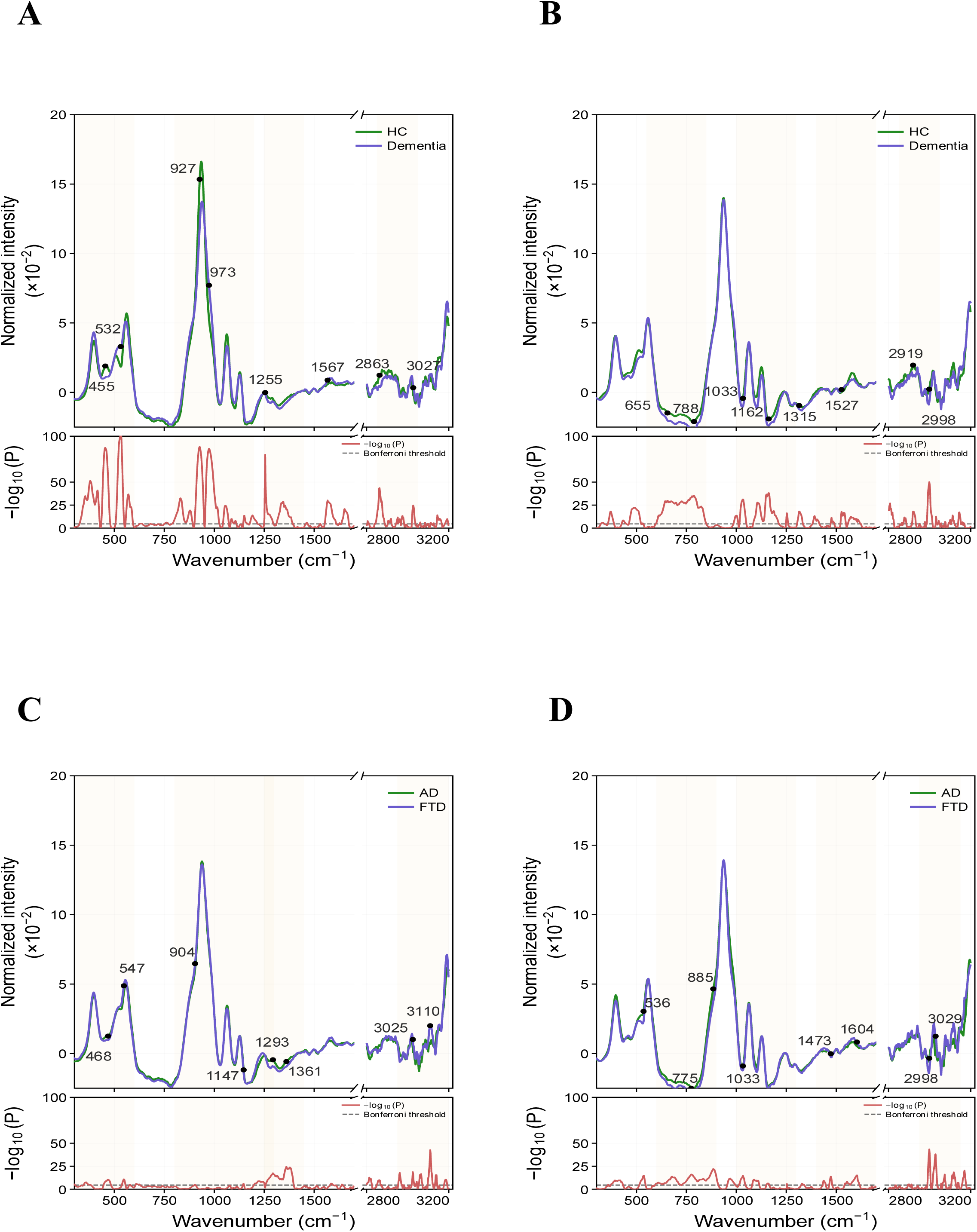
Sex-stratified Raman spectral profiles and wavenumber-wise statistical comparisons among diagnostic groups. **A**, Female HC vs participants with dementia. **B**, Male HC vs participants with dementia. **C**, Female participants with AD vs FTD. **D**, Male participants with AD vs FTD. Upper plots show mean normalized Raman spectra (×10^-2^) across the fingerprint region (300–1700 cm^-1^) and high-wavenumber region (2800–3200 cm^-1^), separated by an axis break. Green and blue lines represent the diagnostic groups indicated in each panel. Black markers and numerical labels denote selected Raman bands of interest. Lower plots show −log₁₀(P) values from wavenumber-wise Welch t tests; red lines represent the −log₁₀(P) profiles, and gray dashed lines indicate the Bonferroni-corrected significance threshold. Lightly shaded areas indicate selected spectral regions of interest. HC, cognitively healthy controls; AD, Alzheimer disease; FTD, frontotemporal dementia.

### Sweat EV Raman profiles show promise for dementia and AD/FTD classification

Participant-level machine-learning models were used to evaluate whether sweat EV Raman spectra could distinguish HC from dementia and, secondarily, AD from FTD, including sex-stratified and balanced-cohort analyses (Figure 3A). In the full HC-vs-dementia cohort (n = 88; 26 HC, 37 AD, and 25 FTD), the best-performing model used SNV normalization, PCA, and a linear support vector machine, achieving an AUC of 0.76, balanced accuracy of 0.68, sensitivity of 0.82, and specificity of 0.54 (Figure 3B–D). Improved performance was observed in a matched balanced cohort comprising 25 HC and 25 participants with dementia (12 AD and 13 FTD), with an AUC of 0.83, balanced accuracy of 0.80, sensitivity of 0.80, and specificity of 0.80 (Figure 3B–D).

**Figure 3.**
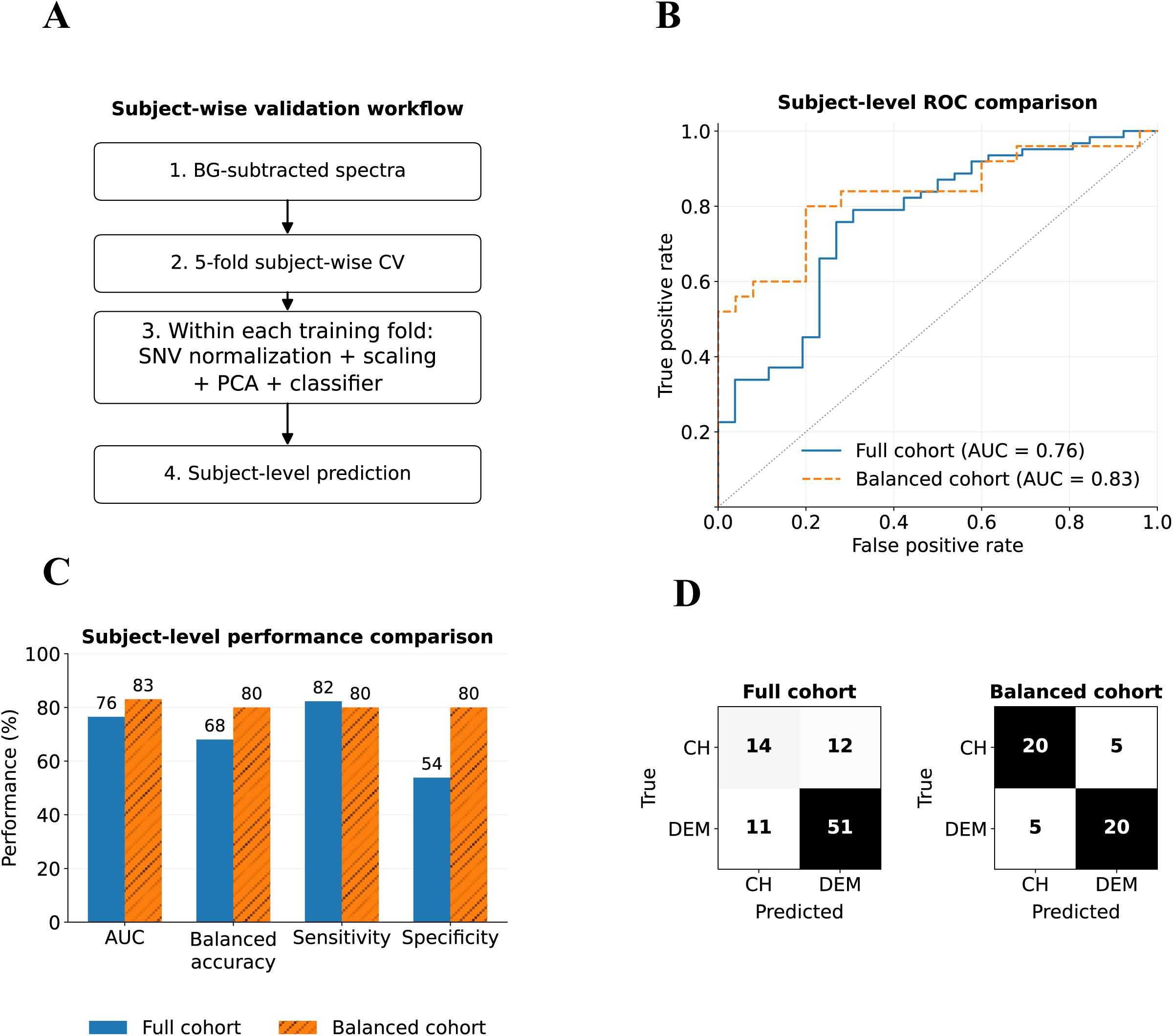
Participant-level machine-learning classification of HC vs dementia using sweat EV Raman spectra. **A** Schematic overview of the classification workflow using background-subtracted Raman spectra, stratified group cross-validation with participants used as the grouping variable, standard normal variate (SNV) normalization, standard scaling, principal component analysis (PCA), classifier fitting, and participant-level prediction. **B,** Receiver operating characteristic (ROC) curves for the full HC-vs-dementia cohort (n = 88) and the matched balanced cohort (n = 50), with area under the ROC curve (AUC) values of 0.76 and 0.83, respectively. **C,** Participant-level classification performance for the full and balanced cohorts, including AUC, balanced accuracy, sensitivity, and specificity. Sensitivity represents dementia detection, and specificity represents correct identification of HC participants. **D,** Confusion matrices showing participant-level predictions for the full and balanced cohorts. Rows indicate true diagnostic groups and columns indicate predicted groups. AUC, area under the receiver operating characteristic curve; EV, extracellular vesicle; HC, cognitively healthy controls; PCA, principal component analysis; ROC, receiver operating characteristic; SNV, standard normal variate. HC, cognitively healthy controls; AD, Alzheimer disease; FTD, frontotemporal dementia.

Sex-specific analyses yielded an AUC of 0.91 in female participants (n = 49; balanced accuracy, 0.82; sensitivity, 0.91; specificity, 0.72) and 0.85 in male participants (n = 39; balanced accuracy, 0.63; sensitivity, 0.93; specificity, 0.33). In the matched balanced analyses, AUCs were 0.89 in females and 0.67 in males, and balanced accuracies were 0.77 and 0.70, respectively (eFigure 7 in the Supplement). Discrimination between AD and FTD was modest. In the overall dementia cohort (n = 62; 37 AD and 25 FTD), the best-performing model achieved an AUC of 0.73, balanced accuracy of 0.68, sensitivity of 0.68, and specificity of 0.68. Similar analyses in the sex-stratified cohorts yielded AUCs of 0.57 in females and 0.78 in males (eFigure 8 in the Supplement). Overall, sweat EV Raman spectra showed stronger classification performance for HC versus dementia than for AD versus FTD.

## Discussion

This study provides proof-of-concept evidence that sweat EVs contain molecular signatures associated with neurodegenerative dementia that can be captured using conventional Raman spectroscopy. AD and FTD were selected because of their clinical relevance and the unmet need for biomarkers to improve diagnostic accuracy, particularly in FTD, where validated minimally invasive disease-specific biomarkers remain limited. Importantly, our findings demonstrate that patients with FTD patients had higher mean sweat EV concentrations than both patients with AD and HC participants, but the difference was significant only between FTD and AD. This difference suggests that altered EV release may reflect disease-specific pathophysiological processes and potentially distinguish these dementia subtypes. This finding is consistent with evidence that neurodegenerative pathology can alter EV biogenesis and cargo composition and that EV dysregulation is associated with cellular stress, neuroinflammation, and metabolic dysfunction ^33,39–44^.

To our knowledge, this is the first study to profile sweat EV Raman signatures in AD, FTD, and HC groups. Across the 300–3200 cm^-1^ spectral range, we identified distinct Raman features that differed between HC and dementia and between AD and FTD. Previous studies have applied Raman spectroscopy to characterize neurodegenerative diseases using brain tissue, CSF, serum, plasma, and whole blood^45–51^. Notably, several differences extended into the high-wavenumber region (approximately 2850–3200 cm^-1^), which comprises C-H stretching vibrations primarily associated with lipids and proteins^52^. These findings may reflect alterations in EV molecular composition and are consistent with previous Raman studies reporting neurodegeneration-associated changes in EV biochemical profiles, particularly in protein and lipid-related spectral features^53,54^. However, molecular assignments based on Raman spectra alone remain tentative and should be validated using complementary proteomic, lipidomic, or metabolomic approaches. The identified Raman bands were therefore interpreted in relation to established biochemical processes implicated in dementia, including protein alterations, lipid metabolism, oxidative stress, and metabolic dysfunction (Table 2).

**Table 2.** Tentative Biomolecular Assignments of Selected Raman Bands in Sweat EVs. Tentative biomolecular assignments of selected Raman wavenumber positions detected in sweat EV spectra. Raman shifts are reported in cm^-1^ and grouped when nearby positions correspond to the same broad vibrational region. Assignments were based on published Raman and EV literature and should be interpreted cautiously because Raman bands may contain overlapping contributions from proteins, lipids, nucleic acids, carbohydrates, and other EV-associated biomolecules. DNA, deoxyribonucleic acid; EV, extracellular vesicle; Phe, phenylalanine; RNA, ribonucleic acid; Trp, tryptophan; Tyr, tyrosine.

| **Biomolecules assignments** | **Molecular vibration** | **Raman Shift (cm<sup>-1</sup>)** | **References** |
| --- | --- | --- | --- |
| **Lipids/Proteins** | CH<sub>2</sub>/CH<sub>3</sub> C–H stretching | 2863, 2919, 2996–2998 | 55,56 |
| **Lipids/Proteins** | =C–H / aromatic C–H stretching | 3025–3029, 3110 | 55–57 |
| **Proteins/ nucleic acids** | Phe/cytosine aromatic ring vibration | 1604 | 58,59 |
| **Proteins / lipids** | Amide I / C=C stretching | 1639 | 57,59 |
| **Protein / nucleic acids** | Aromatic/purine ring vibration | 1585 | 57,60 |
| **Protein** | Trp / purine ring vibration | 1567, 1573 | 60,61 |
| **Aromatic residues / unsaturated lipids** | C=C stretching | 1527 | 62,63 |
| **Lipids / proteins** | CH<sub>2</sub> bending / deformation | 1473 | 59,64 |
| **Proteins / nucleic acids / carbohydrates/lipids** | CH/CH<sub>3</sub> deformation; base/carbohydrate/phospholipid-related vibrations | 1315, 1324, 1345, 1361, 1374, 1384 | 56,60 |
| **Proteins / nucleic acids** | Amide III / DNA ring breathing | 1293 | 65 |
| **Lipids** | CH<sub>2</sub> twisting | 1255 | 65 |
| **Proteins / lipids** | Protein/lipid constituent bands; C–C/C–N stretching | 1147, 1162 | 62 |
| **Nucleic acids** | Phosphodiester groups in nucleic acids | 1079 | 61 |
| **Proteins / lipids** | C–H in-plane vibration; protein–lipid band | 1033 | 60,62 |
| **Carbohydrates / proteins-lipids** | Carbohydrate-associated vibration / C–H in-plane contribution | 1025 | 66 |
| **Proteins / lipids / nucleic acids** | Protein aromatic amino-acid; C–C backbone / lipid-associated vibrations | 885, 925–927, 973, 993 | 56,61,62 |
| **Carbohydrates / EV glycocalyx** | Carbohydrate-originating vibration | 904 | 67 |
| **Nucleic acids** | DNA/RNA base ring breathing; phosphate/backbone contribution | 782, 788 | 56,61 |
| **Proteins** | Tryptophan / aromatic amino-acid band | 775 | 56,60,68 |
| **Proteins** | Tyrosine-related vibration | 677 | 69 |
| **Proteins** | Phenylalanine ring deformation | 655 | 70 |
| **Proteins / lipids** | S–S stretching; cholesterol/cholesterol-ester lipid band | 532, 534–537, 547 | 56,60,71 |
| **Unassigned / low-confidence** | Low-wavenumber skeletal or metal–ligand vibration | 453, 455, 468 | 72 |

Machine-learning analysis showed that sweat EV Raman spectra contained information associated with dementia status. Classification of HC and participants with dementia achieved moderate-to-good-performance, with AUCs of 0.76 in the full cohort and 0.83 in the balanced cohort. The improvement in classification performance after subject-level balancing suggests that the observed signal was not driven solely by class imbalance. Previous Raman studies using plasma, serum, and CSF have reported moderate-to-high diagnostic performance for AD and related dementias^45–47,49^. Our findings extend these observations to sweat EVs, providing a fully non-invasive sample source that captures dementia-associated molecular alterations.

In contrast, discrimination between AD and FTD was modest, with the best-performing model achieved an AUC of 0.73 in the full cohort. Performance differed in sex-stratified analysis, with AUCs of 0.57 in females and 0.78 in males. These findings differ from those reported for pathology-proximal biomarkers, including CSF tau/Ab, plasma p-tau217, and CSF proteomic profiles, which have demonstrated substantially stronger discrimination between AD and FTD^62–64^. The weaker performance observed in the present study may reflect the indirect peripheral nature and greater biological variability of sweat EVs. Despite the limited classification accuracy, several Raman bands differed between AD and FTD, including features near 782, 1374, 1604, 3027, and 3110 cm^-1^ (eTable 2 in the Supplement). These findings suggest that subtype-related molecular variation is present within sweat EVs, although the signal was insufficient for robust participant-level classification in the current cohort. Larger independent cohorts, integration with established biomarkers, and more sensitive Raman approaches will be needed to determine whether sweat EV Raman profiling can contribute to dementia subtype stratification.

Overall, these findings extend Raman-based dementia profiling from blood and CSF to a fully non-invasive sweat EV platform. Although validation in larger independent cohorts is required, the maintained AUC after balancing supports the potential of sweat EV Raman spectroscopy as a complementary approach for dementia characterization and, ultimately, screening or monitoring.

### Data Availability Statement

The data sets generated and/or analysed during the current study are available from the corresponding author on request.

### Institutional Review Board Statement

The study was conducted in accordance with the Declaration of Helsinki and has been approved by the Northern Ostrobothnia Hospital District Ethics committee (EETMK: 210/2001).

### Informed Consent Statement

Informed consent was obtained from all participants involved in the study.

## Authors Contributions

A.U.D performed the Raman spectroscopy measurements and analysis of sweat EVs. A.U.D also prepared the final figures, conducted the classification analyses, and contributed to the preparation of the first draft of the manuscript. S.Sh. performed the isolation and characterization of sweat EVs, including NTA, prepared the samples for EM analysis, and contributed to the writing of the first draft of the manuscript. E.A. also participated in the isolation and characterization of EVs. M.M. supported the optimization of the Raman spectroscopy measurements, provided continuous guidance to A.U.D throughout the project, and contributed to the Python-based data analysis. S.S. contributed to the AI-based analyses and critically revised the manuscript. J.K. conducted the recruitment, clinical assessments and characterization of the study participants, contributed to manuscript preparation and conceptualization of the work, and critically revised the manuscript. N.A. Conceptualized and designed the study; defined the scientific scope; supervised the research and students; contributed to the drafting, review, and revision of the manuscript; served as corresponding author; and served as guarantor, taking responsibility for the overall integrity of the work.

## Acknowledgements

We sincerely thank Pirkko Holappa for her invaluable assistance with the recruitment of participants, collection of biological samples, and administration of the cognitive assessments. We also thank Ilkka Miinalainen for his support with the electron microscopy (EM) analysis. Finally, we are grateful to all the participants who generously contributed to this study.

## References

1. Dementia. Accessed June 18, 2025. https://www.who.int/news-room/fact-sheets/detail/dementia

2. ADI - WHO launches global status report on public health response to dementia. Accessed June 18, 2025. https://www.alzint.org/news-events/news/who-launches-its-global-status-report-on-the-public-health-response-to-dementia/

3. Estimation of the global prevalence of dementia in 2019 and forecasted prevalence in 2050: an analysis for the Global Burden of Disease Study 2019. Lancet Public Health. 2022;7(2):e105–e125. doi:10.1016/S2468-2667(21)00249-8

4. Saloner R, Staffaroni AM, Dammer EB, et al. Large-scale network analysis of the cerebrospinal fluid proteome identifies molecular signatures of frontotemporal lobar degeneration. Nat Aging. Published online May 16, 2025:1–16. doi:10.1038/s43587-025-00878-2

5. Vermunt L, Otte M, Verberk IMW, et al. Age- and disease-specific reference values for neurofilament light presented in an online interactive support interface. Ann Clin Transl Neurol. 2022;9(11):1832–1837. doi:10.1002/acn3.51676

6. Taddei RN, Duff KE. Synapse vulnerability and resilience across the clinical spectrum of dementias. Nat Rev Neurol. Published online May 22, 2025:1–17. doi:10.1038/s41582-025-01094-7

7. Baiardi S, Quadalti C, Mammana A, et al. Diagnostic value of plasma p-tau181, NfL, and GFAP in a clinical setting cohort of prevalent neurodegenerative dementias. Alzheimers Res Ther. 2022;14:153. doi:10.1186/s13195-022-01093-6

8. Gómez-Tortosa E, Agüero-Rabes P, Ruiz-González A, et al. Plasma Biomarkers in the Distinction of Alzheimer’s Disease and Frontotemporal Dementia. Int J Mol Sci. 2025;26(3):1231. doi:10.3390/ijms26031231

9. Liscic RM, Storandt M, Cairns NJ, Morris JC. Clinical and psychometric distinction of frontotemporal and Alzheimer dementias. Arch Neurol. 2007;64(4):535–540. doi:10.1001/archneur.64.4.535

10. Padovani A, Premi E, Pilotto A, et al. Overlap between frontotemporal dementia and Alzheimer’s disease: cerebrospinal fluid pattern and neuroimaging study. J Alzheimers Dis. 2013;36(1):49–55. doi:10.3233/JAD-121969

11. Bright F, Werry EL, Dobson-Stone C, et al. Neuroinflammation in frontotemporal dementia. Nat Rev Neurol. 2019;15(9):540–555. doi:10.1038/s41582-019-0231-z

12. del Campo M, Zetterberg H, Gandy S, et al. New developments of biofluid-based biomarkers for routine diagnosis and disease trajectories in frontotemporal dementia. Alzheimers Dement. 2022;18(11):2292–2307. doi:10.1002/alz.12643

13. McDade E, Cummings JL, Dhadda S, et al. Lecanemab in patients with early Alzheimer’s disease: detailed results on biomarker, cognitive, and clinical effects from the randomized and open-label extension of the phase 2 proof-of-concept study. Alz Res Therapy. 2022;14(1):191. doi:10.1186/s13195-022-01124-2

14. Dyck CH van, Swanson CJ, Aisen P, et al. Lecanemab in Early Alzheimer’s Disease. New England Journal of Medicine. 2023;388(1):9–21. doi:10.1056/NEJMoa2212948

15. Sims JR, Zimmer JA, Evans CD, et al. Donanemab in Early Symptomatic Alzheimer Disease. JAMA. 2023;330(6):512–527. doi:10.1001/jama.2023.13239

16. Zeng B, Tang C, Wang J, Yang Q, Ren Q, Liu X. Pharmacologic and Nutritional Interventions for Early Alzheimer’s Disease: A Systematic Review and Network Meta-Analysis of Randomized Controlled Trials. J Alzheimers Dis. 99(4):1173–1186. doi:10.3233/JAD-240161

17. Ferretti MT, Iulita MF, Cavedo E, et al. Sex differences in Alzheimer disease — the gateway to precision medicine. Nat Rev Neurol. 2018;14(8):457–469. doi:10.1038/s41582-018-0032-9

18. Gregory S, Bridgeman K, Darwin H, et al. Associations of estrogen with modifiable and non-modifiable risk factors for dementia: A narrative review. Alzheimer’s & Dementia. 2025;21(11):e70873. doi:10.1002/alz.70873

19. Lopez-Lee C, Torres ERS, Carling G, Gan L. Mechanisms of sex differences in Alzheimer’s disease. Neuron. 2024;112(8):1208–1221. doi:10.1016/j.neuron.2024.01.024

20. Price BR, Walker KA, Eissman JM, et al. Sex differences and the role of estrogens in the immunological underpinnings of Alzheimer’s disease. Alzheimer’s & Dementia: Translational Research & Clinical Interventions. 2025;11(3):e70139. doi:10.1002/trc2.70139

21. Eikelboom WS, Pan M, Ossenkoppele R, et al. Sex differences in neuropsychiatric symptoms in Alzheimer’s disease dementia: a meta-analysis. Alz Res Therapy. 2022;14(1):48. doi:10.1186/s13195-022-00991-z

22. Silvestri C, Almici V, Libri I, et al. Sex Differences in the Severity and Progression of Neuropsychiatric Symptoms Across Different Dementia Types. Neurology Clinical Practice. 2024;14(3):e200299. doi:10.1212/CPJ.0000000000200299

23. Liesinger AM, Graff-Radford NR, Duara R, et al. Sex and age interact to determine clinicopathologic differences in Alzheimer’s disease. Acta Neuropathol. 2018;136(6):873–885. doi:10.1007/s00401-018-1908-x

24. Hanna K, Krzoska E, Shaaban AM, Muirhead D, Abu-Eid R, Speirs V. Raman spectroscopy: current applications in breast cancer diagnosis, challenges and future prospects. Br J Cancer. 2022;126(8):1125–1139. doi:10.1038/s41416-021-01659-5

25. Wang Y, Fang L, Wang Y, Xiong Z. Current Trends of Raman Spectroscopy in Clinic Settings: Opportunities and Challenges. Advanced Science. 2024;11(7):2300668. doi:10.1002/advs.202300668

26. Han XX, Rodriguez RS, Haynes CL, Ozaki Y, Zhao B. Surface-enhanced Raman spectroscopy. Nat Rev Methods Primers. 2022;1(1):1–17. doi:10.1038/s43586-021-00083-6

27. Ember KJI, Hoeve MA, McAughtrie SL, et al. Raman spectroscopy and regenerative medicine: a review. npj Regen Med. 2017;2(1):1–10. doi:10.1038/s41536-017-0014-3

28. Rahat ST, Mäkelä M, Nasserinejad M, et al. Clinical-Grade Patches as a Medium for Enrichment of Sweat-Extracellular Vesicles and Facilitating Their Metabolic Analysis. Int J Mol Sci. 2023;24(8):7507. doi:10.3390/ijms24087507

29. Ali N, Rahat ST, Mäkelä M, et al. Metabolic patterns of sweat-extracellular vesicles during exercise and recovery states using clinical grade patches. Front Physiol. 2023;14:1295852. doi:10.3389/fphys.2023.1295852

30. Zhyvolozhnyi A, Samoylenko A, Bart G, et al. Enrichment of sweat-derived extracellular vesicles of human and bacterial origin for biomarker identification. Nanotheranostics. 2024;8(1):48–63. doi:10.7150/ntno.87822

31. Kim J, Yang JD, Agopian VG, Zhu Y, Tseng HR, You S. Computational frameworks for enhanced extracellular vesicle biomarker discovery. Exp Mol Med. 2026;58(1):73–81. doi:10.1038/s12276-025-01622-x

32. Xiao J, Sluijter JPG. Extracellular vesicles in cardiovascular homeostasis and disease: potential role in diagnosis and therapy. Nat Rev Cardiol. 2025;22(11):883–895. doi:10.1038/s41569-025-01141-2

33. Manolopoulos A, Yao PJ, Kapogiannis D. Extracellular vesicles: translational research and applications in neurology. Nat Rev Neurol. 2025;21(5):265–282. doi:10.1038/s41582-025-01080-z

34. Dubois B, Villain N, Frisoni GB, et al. Clinical diagnosis of Alzheimer’s disease: recommendations of the International Working Group. The Lancet Neurology. 2021;20(6):484–496. doi:10.1016/S1474-4422(21)00066-1

35. Rascovsky K, Hodges JR, Knopman D, et al. Sensitivity of revised diagnostic criteria for the behavioural variant of frontotemporal dementia. Brain. 2011;134(Pt 9):2456–2477. doi:10.1093/brain/awr179

36. Gorno-Tempini ML, Hillis AE, Weintraub S, et al. Classification of primary progressive aphasia and its variants. Neurology. 2011;76(11):1006–1014. doi:10.1212/WNL.0b013e31821103e6

37. Moms JC, Heyman A, Mohs RC, et al. The Consortium to Establish a Registry for Alzheimer’s Disease (CERAD). Part I. Clinical and neuropsychological assesment of Alzheimer’s disease. Neurology. 1989;39(9):1159–1159. doi:10.1212/WNL.39.9.1159

38. Rahat ST, Miinalainen I, Nieminen AI, Vainio S, Ali N. Heat stress alters metabolic pathways and nitric oxide signaling in keratinocytes under hyperglycemia. Journal of Diabetes Investigation. 2026;17(4):608–621. doi:10.1111/jdi.70256

39. Rydland A, Yran ES, Nyman TA, et al. Exploring differences in protein cargo of extracellular vesicles from ME/CFS patient plasma compared to healthy controls. Biochem Biophys Rep. 2026;47:102679. doi:10.1016/j.bbrep.2026.102679

40. Kalluri R. The biology and function of extracellular vesicles in immune response and immunity. Immunity. 2024;57(8):1752–1768. doi:10.1016/j.immuni.2024.07.009

41. Spiers JG, Vassileff N, Hill AF. Neuroinflammatory Modulation of Extracellular Vesicle Biogenesis and Cargo Loading. Neuromol Med. 2022;24(4):385–391. doi:10.1007/s12017-022-08704-3

42. Thompson AG, Gray E, Heman-Ackah SM, et al. Extracellular vesicles in neurodegenerative disease — pathogenesis to biomarkers. Nat Rev Neurol. 2016;12(6):346–357. doi:10.1038/nrneurol.2016.68

43. Wiersema AF, Rennenberg A, Smith G, Varderidou-Minasian S, Pasterkamp RJ. Shared and distinct changes in the molecular cargo of extracellular vesicles in different neurodegenerative diseases. Cell Mol Life Sci. 2024;81(1):479. doi:10.1007/s00018-024-05522-7

44. Wilson MR, Satapathy S, Vendruscolo M. Extracellular protein homeostasis in neurodegenerative diseases. Nat Rev Neurol. 2023;19(4):235–245. doi:10.1038/s41582-023-00786-2

45. Carota AG, Campanella B, Del Carratore R, Bongioanni P, Giannelli R, Legnaioli S. Raman spectroscopy and multivariate analysis as potential tool to follow Alzheimer’s disease progression. Anal Bioanal Chem. 2022;414(16):4667–4675. doi:10.1007/s00216-022-04087-3

46. Devitt G, Michopoulou SK, Kadalayil L, et al. Classification of Alzheimer’s disease in a mixed clinical cohort using biofluid Raman spectroscopy. Alz Res Therapy. 2025;17(1):228. doi:10.1186/s13195-025-01879-4

47. Conti F, Banchelli M, Bessi V, et al. Alzheimer Disease Detection from Raman Spectroscopy of the Cerebrospinal Fluid via Topological Machine Learning. Engineering Proceedings. 2023;51(1):14. doi:10.3390/engproc2023051014

48. Lochocki B, Boon BDC, Verheul SR, et al. Multimodal, label-free fluorescence and Raman imaging of amyloid deposits in snap-frozen Alzheimer’s disease human brain tissue. Commun Biol. 2021;4(1):474. doi:10.1038/s42003-021-01981-x

49. Paraskevaidi M, Morais CLM, Halliwell DE, et al. Raman Spectroscopy to Diagnose Alzheimer’s Disease and Dementia with Lewy Bodies in Blood. ACS Chem Neurosci. 2018;9(11):2786–2794. doi:10.1021/acschemneuro.8b00198

50. Habartová L, Hrubešová K, Syslová K, et al. Blood-based molecular signature of Alzheimer’s disease *via* spectroscopy and metabolomics. Clinical Biochemistry. 2019;72:58–63. doi:10.1016/j.clinbiochem.2019.04.004

51. Lopez E, Etxebarria-Elezgarai J, García-Sebastián M, et al. Unlocking Preclinical Alzheimer’s: A Multi-Year Label-Free In Vitro Raman Spectroscopy Study Empowered by Chemometrics. International Journal of Molecular Sciences. 2024;25(9):4737. doi:10.3390/ijms25094737

52. Gualerzi A, Kooijmans SAA, Niada S, et al. Raman spectroscopy as a quick tool to assess purity of extracellular vesicle preparations and predict their functionality. J Extracell Vesicles. 2019;8(1):1568780. doi:10.1080/20013078.2019.1568780

53. Gualerzi A, Picciolini S, Carlomagno C, et al. Raman profiling of circulating extracellular vesicles for the stratification of Parkinson’s patients. Nanomedicine: Nanotechnology, Biology and Medicine. 2019;22:102097. doi:10.1016/j.nano.2019.102097

54. Imanbekova M, Suarasan S, Rojalin T, et al. Identification of amyloid beta in small extracellular vesicles via Raman spectroscopy. Nanoscale Adv. 3(14):4119–4132. doi:10.1039/d1na00330e

55. Buccini L, Proietti A, Penna GL, et al. Toward the nanoscale chemical and physical probing of milk-derived extracellular vesicles using Raman and tip-enhanced Raman spectroscopy. Nanoscale. 2024;16(16):8132–8142. doi:10.1039/D4NR00845F

56. Gualerzi A, Niada S, Giannasi C, et al. Raman spectroscopy uncovers biochemical tissue-related features of extracellular vesicles from mesenchymal stromal cells. Sci Rep. 2017;7(1):9820. doi:10.1038/s41598-017-10448-1

57. Adar F. Interpretation of Raman Spectrum of Proteins | Spectroscopy Online. July 20, 2026. Accessed July 20, 2026. https://www.spectroscopyonline.com/view/interpretation-of-raman-spectrum-of-proteins

58. Morasso CF, Sproviero D, Mimmi MC, et al. Raman spectroscopy reveals biochemical differences in plasma derived extracellular vesicles from sporadic Amyotrophic Lateral Sclerosis patients. Nanomedicine: Nanotechnology, Biology and Medicine. 2020;29:102249. doi:10.1016/j.nano.2020.102249

59. Koster HJ, Rojalin T, Powell A, et al. Surface enhanced Raman scattering of extracellular vesicles for cancer diagnostics despite isolation dependent lipoprotein contamination. Nanoscale. 13(35):14760–14776. doi:10.1039/d1nr03334d

60. Zhang P, Wang L, Fang Y, Zheng D, Lin T, Wang H. Label-Free Exosomal Detection and Classification in Rapid Discriminating Different Cancer Types Based on Specific Raman Phenotypes and Multivariate Statistical Analysis. Molecules. 2019;24(16):2947. doi:10.3390/molecules24162947

61. Zhang H, Silva AC, Zhang W, Rutigliano H, Zhou A. Raman Spectroscopy characterization extracellular vesicles from bovine placenta and peripheral blood mononuclear cells. PLOS ONE. 2020;15(7):e0235214. doi:10.1371/journal.pone.0235214

62. Shin H, Choi BH, Shim O, et al. Single test-based diagnosis of multiple cancer types using Exosome-SERS-AI for early stage cancers. Nat Commun. 2023;14(1):1644. doi:10.1038/s41467-023-37403-1

63. Ćulum NM, Cooper TT, Bell GI, Hess DA, Lagugné-Labarthet F. Characterization of extracellular vesicles derived from mesenchymal stromal cells by surface-enhanced Raman spectroscopy. Anal Bioanal Chem. 2021;413(20):5013–5024. doi:10.1007/s00216-021-03464-8

64. Buchan E, Rickard JJS, Thomas MR, Oppenheimer PG. Advanced Multipurpose Spectroscopic Nanobio-Device for Concurrent Lab-on-a-Chip Label-Free Separation and Detection of Extracellular Vesicles as Key-Biomarkers for Point-of-Care Cardiovascular Disease Diagnostics. Adv Healthc Mater. 2025;14(21):e2500122. doi:10.1002/adhm.202500122

65. Penders J, Nagelkerke A, Cunnane EM, et al. Single Particle Automated Raman Trapping Analysis of Breast Cancer Cell-Derived Extracellular Vesicles as Cancer Biomarkers. ACS Nano. 2021;15(11):18192–18205. doi:10.1021/acsnano.1c07075

66. Parlatan U, Ozen MO, Kecoglu I, et al. Label-Free Identification of Exosomes using Raman Spectroscopy and Machine Learning. Small. 2023;19(9):2205519. doi:10.1002/smll.202205519

67. Rojalin T, Koster HJ, Liu J, et al. Hybrid Nanoplasmonic Porous Biomaterial Scaffold for Liquid Biopsy Diagnostics Using Extracellular Vesicles. ACS Sens. 2020;5(9):2820–2833. doi:10.1021/acssensors.0c00953

68. Kamińska A, Roman M, Wróbel A, et al. Raman spectroscopy of urinary extracellular vesicles to stratify patients with chronic kidney disease in type 2 diabetes. Nanomedicine: Nanotechnology, Biology and Medicine. 2022;39:102468. doi:10.1016/j.nano.2021.102468

69. Hao N, Wang Z, Liu P, et al. Acoustofluidic multimodal diagnostic system for Alzheimer’s disease. Biosensors and Bioelectronics. 2022;196:113730. doi:10.1016/j.bios.2021.113730

70. Tan B, Venkatakrishnan K, GANESH S, Ravindra HR. Apparatus and method for early cancer detection and cancer prognosis using a nanosensor with raman spectroscopy. Published online August 25, 2022:WO2022174346A1. Accessed July 11, 2025. https://patents.google.com/patent/WO2022174346A1/en

71. McAvan BS, Bowsher LA, Powell T, et al. Raman Spectroscopy to Monitor Post-Translational Modifications and Degradation in Monoclonal Antibody Therapeutics. Anal Chem. 2020;92(15):10381–10389. doi:10.1021/acs.analchem.0c00627

72. Lopes J, Correia M, Martins I, et al. FTIR and Raman Spectroscopy Applied to Dementia Diagnosis Through Analysis of Biological Fluids. Journal of Alzheimer’s Disease. 2016;52(3):801–812. doi:10.3233/JAD-151163

