## Supplementary material for "Decoding Dementia from Sweat: A Proof-of-Concept Label-Free Raman Fingerprinting of EVs": Online Supplement material


**eMethods**

**eReferences**

**eTable 1.** Raman Intensities at Selected Wavenumbers in Cognitively Healthy Controls and Participants With Dementia

**eTable 2.** Raman Intensities at Selected Wavenumbers in AD and FTD

**eTable 3.** Sex-Stratified Raman Intensities at Selected Wavenumbers in HC and Dementia Groups

**eTable 4.** Sex-Stratified Raman Intensities at Selected Wavenumbers in AD and FTD

**eFigure 1.** Raman Spectral Preprocessing Workflow for Sweat EV Samples

**eFigure 2.** Characterization of Sweat EVs by Nanoparticle Tracking Analysis and Transmission Electron Microscopy

**eFigure 3.** Raman Intensity Distributions at Selected Wavenumbers in Cognitively Healthy Controls and Participants With Dementia

**eFigure 4.** Raman Intensity Distributions at Selected Wavenumbers in AD and FTD

**eFigure 5.** Sex-Stratified Raman Intensity Distributions in Cognitively Healthy Controls and Participants With Dementia

**eFigure 6.** Sex-Stratified Raman Intensity Distributions in AD and FTD

**eFigure 7.** Sex-Stratified HC vs Dementia Classification in Full and Balanced Cohorts

**eFigure 8.** AD vs FTD Classification Overall and by Sex

### eMethods

#### Sweat Patch Collection

During each visit, two biological samples were collected from participants, including natural sweat captured using clinical grade sweat patches (Sorbact® 10 × 10 cm [cat. no. 98501] and Mesoft® 5 × 5 cm [Pharmacy]). Patches were pre-assembled by affixing an adhesive layer, Tegaderm™ transparent film, to the absorbent patch material. This transparent film served to secure the patch to the skin and prevent sweat evaporation. The prepared patches were then applied to the upper side of participants' arm. After 45 minutes of adherence, the patches were carefully removed. The adhesive film was discarded, and the collected sweat-containing absorbent material was then stored at −70 °C until further analysis.

#### Extracellular Vesicle Isolation From Sweat Patches

In brief, after collection, the Sorbact® (10 × 10 cm [cat. no. 98501) or Mesoft® (5 × 5 cm, available in Pharmacy) dressing was opened, and the superabsorbent layer was dissolved in 15 mL (1×) phosphate-buffered saline (PBS). The content was then filtered through 0.8-µm Minisart® NML syringe filters (Sartorius). The filtrate was subjected to ultracentrifugation of 100,000 × g for 22 h in a Sorvall WX Ultra 90 ultracentrifuge using a Sorvall AH-629 rotor (VWR, Thermo Electrom Corporation). After careful removal of the supernatant, the pellet was resuspended and eluted in 60 µL of 1× PBS and stored at −70 °C for subsequent analysis.

#### Nanoparticle Tracking Analysis

Nanoparticle tracking analysis (NTA) was performed using a NanoSight NS300 (NanoSight Ltd, Amesbury, UK) equipped with a 405-nm laser. At least four 60 s videos were recorded for each sample with camera level 14 and detection threshold set at 5. Temperature was monitored throughout the measurements. Videos recorded for each sample were analyzed with NTA software version 3 to determine the concentration and size of measured particles with the corresponding standard error. For analysis, auto settings were used for blur, minimum track length and minimum expected particle size. Double-distilled $H_2O$ was used to dilute the sample.

#### Transmission Electron Microscopy and Negative Staining

Isolated EVs were settled on Formvar carbonated copper grid (glow-discharged). Following fixation with 1% glutaraldehyde, the grid was washed with distilled water and stained with neutral 2% Uranyl acetate (UA). Afterwards, the grid was coated with 2% methylcellulose-UA solution and following 10 min incubation the excess fluid was carefully removed, and grids were air dried. EVs were visualized using a JEOL JEM-120i 120 kV transmission electron microscope and images were taken with a Veleta charge-coupled device (CCD) camera and Item software (Olympus Soft Imaging Solutions GMBH, Munster, Germany).

#### Raman Spectroscopy Sample Preparation and Acquisition

Prior to Raman measurement, 5 µL of each sweat EV sample was deposited onto an aluminium mirror substrate and allowed to air-dry for at least 45 minutes. Raman spectra were acquired using a DXR2xi Raman Imaging Microscope (Thermo Fisher Scientific) operated with OMNICxi Raman Imaging Software. Measurements were performed using a 785-nm excitation laser with a full-range grating covering 50–3200 $cm^{-1}$, a 50× objective, and a 50-µm slit aperture. Each acquisition used a 1-second exposure averaged over 10 scans, with laser power between 26.5 and 27.0 mW. Spectral maps were acquired using pixel sizes ranging from 1.6 to 18 µm, depending on the scanned area. Approximately 160–200 spectra were collected from each sample, predominantly from the peripheral regions of the air-dried droplet.

#### Raman Spectral Preprocessing

Raw Raman data were imported and converted into spectral arrays. Spectra were quality-filtered using the high-wavenumber region (2000–3200 $cm^{-1}$), including cosmic-ray despiking[1] and signal-to-noise–based spectral selection. Spectra passing quality control were clipped or refitted to a common wavenumber range of 300–3200 $cm^{-1}$.

Spectral smoothing was performed using a Savitzky-Golay filter with a polynomial order of 3 and a window size of 25 points.[2] Baseline correction was then performed using Whittaker asymmetric least-squares baseline correction.[3] Background spectra acquired from the aluminium mirror substrate underwent the same core preprocessing steps as the sample spectra.

For normalization, the maximum intensity within the 1200–1300 $cm^{-1}$ region of the preprocessed background spectra was used as the scaling reference. Each sample spectrum was scaled using a multiplier calculated as the ratio of the background reference intensity to the corresponding maximum sample intensity within the same region. Baseline correction was performed before normalization.[4] Following normalization, the mean preprocessed background spectrum was subtracted from each sample spectrum to reduce substrate-related background contributions. The resulting background-subtracted spectra were used for downstream statistical and machine-learning analyses. The complete preprocessing workflow is illustrated in eFigure 1 in the Supplement.

**Wavenumber-Wise Statistical Analysis**

Group-level Raman spectral differences were evaluated using the background-subtracted spectra. Comparisons were performed between HC and dementia, between AD and FTD, and separately within female and male participants.

At each wavenumber, normalized Raman intensities were compared between groups using 2-sided Welch independent-samples *t* tests. Welch testing was used because equal numbers of spectra and equal within-group variances were not assumed. To account for multiple wavenumber-wise comparisons, the significance threshold was adjusted using the Bonferroni method across all tested wavenumber points. The resulting *P* values were transformed to $-\log_{10}(P)$ to generate wavenumber-wise statistical significance profiles.

To summarize adjacent significant spectral points for visualization, local maxima exceeding the Bonferroni-corrected significance threshold were identified from each $-\log_{10}(P)$ profile. Within each predefined reporting region, 2 of the strongest significant local maxima separated by at least 25 $cm^{-1}$ were selected for annotation. The reporting regions were 300–800, 800–1200, 1200–1700, and 2850–3200 $cm^{-1}$.

**Visualization of Selected Raman Bands**

Intensity distributions at the selected wavenumbers were visualized using boxplots. All available spectra were included in the boxplot calculations, whereas only a randomly selected subset of individual spectra was displayed as overlaid points to improve visual clarity. Mean (SEM) values and exact spectrum-level Welch *P* values were calculated for the selected wavenumbers.

Because multiple Raman spectra were obtained from each participant, the wavenumber-wise comparisons were treated as exploratory spectrum-level analyses rather than independent participant-level statistical tests. Participant-level discrimination was evaluated separately using the machine-learning procedure described below.

**Machine-Learning Classification: Tasks and Spectral Input**

Machine-learning analyses were performed using background-subtracted sweat extracellular vesicle (EV) Raman spectra.[5] Two binary classification tasks were evaluated. For cognitively healthy controls (HC) vs dementia, HC participants were assigned to class 0 and participants with Alzheimer disease (AD) or frontotemporal dementia (FTD) were assigned to class 1. For AD vs FTD classification, AD participants were assigned to class 0 and FTD participants were assigned to class 1. Both classification tasks were additionally evaluated separately in female and male participants. Before model fitting, Raman spectra were aligned to a common wavenumber axis when individual spectra contained separate wavenumber axes. A common overlapping spectral range was determined and spectra were linearly interpolated onto the shared axis. When the data already contained a single common wavenumber axis, no interpolation was applied. Only the background-subtracted spectral dataset was used for the reported classification analyses.

**Cross-validation and machine-learning pipeline**

Model evaluation was performed using stratified group cross-validation with participants used as the grouping variable. This ensured that all spectra from a given participant were assigned exclusively to either the training or test set within each fold.[6,7] Cross-validation used a maximum of 5 folds with shuffling and a random state of 42. When the number of participants in the smallest class was insufficient for 5-fold cross-validation, the number of folds was reduced accordingly. Within each cross-validation fold, spectra underwent standard normal variate (SNV) normalization, standard scaling, principal component analysis (PCA), and classifier fitting. SNV normalization was applied independently to each spectrum by subtracting its mean intensity and dividing by its standard deviation. Spectra with a standard deviation of 0 were assigned a denominator of 1 for this transformation. Standard scaling was subsequently performed using StandardScaler. PCA used the randomized solver with a random state of 42. Standard scaling and PCA were fitted using the training data within each cross-validation fold and subsequently applied to the corresponding held-out test data. SNV normalization, scaling, PCA, and classifier fitting were implemented using the scikit-learn Pipeline framework. Two linear classifiers were evaluated: logistic regression and a linear support vector classifier. Logistic regression was implemented using the lbfgs solver, balanced class weighting, and a maximum of 3000 iterations. The linear support vector classifier used balanced class weighting, dual='auto', and a maximum of 10 000 iterations.

**Participant-level prediction and performance evaluation**

Classifier decision scores were initially generated for individual spectra in each held-out cross-validation fold. Scores from all spectra belonging to the same participant were then averaged to generate a single participant-level decision score. Because both evaluated classifiers provided decision-function scores, participant-level class predictions were assigned using a decision threshold of 0. Classification performance was calculated at the participant level using area under the receiver operating characteristic curve (AUC), accuracy, balanced accuracy, sensitivity, specificity, precision, and F1 score. For HC-vs-dementia classification, sensitivity represented correct identification of participants with dementia and specificity represented correct identification of HC participants. For AD-vs-FTD classification, sensitivity represented correct identification of FTD and specificity represented correct identification of AD. Performance metrics were calculated within each cross-validation fold, and mean and SD values across folds were generated for each model configuration. Model configurations were ranked primarily according to mean cross-validated AUC. When required for ranking, mean balanced accuracy and then mean accuracy were used as secondary criteria. Confusion matrices and receiver operating characteristic curves were generated from the pooled held-out participant-level predictions. The reported classification performance therefore represents internal participant-grouped cross-validation rather than evaluation in an independent external test cohort.

**Balanced HC-vs-Dementia Sensitivity Analyses**

Additional sensitivity analyses were performed to evaluate the influence of class imbalance on HC-vs-dementia classification. Participant selection was performed before cross-validation, and all spectra belonging to each selected participant were retained. Participant selection used a fixed random state of 42. For the overall HC-vs-dementia balanced analysis, the target class size was 25 participants per group. An equal number of participants with dementia was subsequently selected. Within the dementia group, the selection procedure targeted equal representation of AD and FTD and a female-to-male distribution matching that of the HC group as closely as possible. Candidate dementia subsets were sampled repeatedly, for a maximum of 20 000 iterations, and the subset minimizing the combined deviation from the target diagnostic and sex distributions was retained. Sampling terminated earlier when an exact match to the prespecified targets was identified. For sex-specific HC-vs-dementia balanced analyses, participants were first restricted to the relevant sex and equal numbers of HC and dementia participants were then selected according to the number available in the smaller classification group. All Raman spectra belonging to the selected participants were retained. The same participant-grouped cross-validation and classification procedures described above were subsequently applied.

**Software**

Machine-learning analyses were implemented in Python using NumPy, pandas, and scikit-learn, with matplotlib used for visualization. Classification pipelines were constructed using the scikit-learn Pipeline framework, and grouped cross-validation was implemented using StratifiedGroupKFold.

### References


1. Whitaker DA, Hayes K. A simple algorithm for despiking Raman spectra. *Chemometrics and Intelligent Laboratory Systems*. 2018;179:82-84. doi:10.1016/j.chemolab.2018.06.009
2. Savitzky Abraham, Golay MJE. Smoothing and Differentiation of Data by Simplified Least Squares Procedures. *Anal Chem*. 1964;36(8):1627-1639. doi:10.1021/ac60214a047
3. Eilers P, Boelens H. Baseline Correction with Asymmetric Least Squares Smoothing. *Unpubl Manuscr*. Published online November 21, 2005.
4. Afseth NK, Segtnan VH, Wold JP. Raman Spectra of Biological Samples: A Study of Preprocessing Methods. *Appl Spectrosc, AS*. 2006;60(12):1358-1367.
5. Oancea B, Seciu-Grama AM, Siminea N, et al. Machine Learning for Biomedical Raman Spectroscopy: From Spectral Acquisition to Clinical Translation. *arXiv*. Preprint posted online June 12, 2026:arXiv:2606.14169. doi:10.48550/arXiv.2606.14169
6. Beleites C, Neugebauer U, Bocklitz T, Krafft C, Popp J. Sample size planning for classification models. *Analytica Chimica Acta*. 2013;760:25-33. doi:10.1016/j.aca.2012.11.007
7. Blake N, Gaifulina R, Griffin LD, Bell IM, Thomas GMH. Machine Learning of Raman Spectroscopy Data for Classifying Cancers: A Review of the Recent Literature. *Diagnostics*. 2022;12(6):1491. doi:10.3390/diagnostics12061491

### eTable 1. Raman Intensities at Selected Wavenumbers in Cognitively Healthy Controls and Participants With Dementia

| Wavenumber ($cm^{-1}$) | HC (mean ± SEM) | Dementia (mean ± SEM) | Welch P value |
|---|---|---|---|
| 453 | 0.0165 ± 0.0003 | 0.0110 ± 0.0002 | $1.97 \times 10^{-64}$ |
| 534 | 0.0233 ± 0.0004 | 0.0301 ± 0.0004 | $3.51 \times 10^{-34}$ |
| 925 | 0.1377 ± 0.0010 | 0.1184 ± 0.0008 | $2.86 \times 10^{-51}$ |
| 993 | 0.0339 ± 0.0004 | 0.0437 ± 0.0004 | $3.31 \times 10^{-63}$ |
| 1324 | -0.0099 ± 0.0002 | -0.0123 ± 0.0001 | $7.47 \times 10^{-32}$ |
| 1384 | -0.0017 ± 0.0002 | -0.0037 ± 0.0001 | $1.37 \times 10^{-18}$ |
| 2863 | 0.0108 ± 0.0004 | 0.0046 ± 0.0002 | $1.23 \times 10^{-39}$ |
| 2996 | -0.0009 ± 0.0003 | -0.0060 ± 0.0002 | $2.99 \times 10^{-35}$ |

Abbreviations: HC, cognitively healthy controls; SEM, standard error of the mean.
P values were calculated using 2-sided spectrum-level Welch independent-samples t tests. Because multiple Raman spectra were obtained from each participant, these comparisons represent exploratory spectrum-level analyses.

### eTable 2. Raman Intensities at Selected Wavenumbers in AD and FTD

| Wavenumber ($cm^{-1}$) | AD (mean ± SEM) | FTD (mean ± SEM) | P value |
|---|---|---|---|
| 677 | -0.0179 ± 0.0002 | -0.0205 ± 0.0003 | $1.14 \times 10^{-13}$ |
| 782 | -0.0238 ± 0.0003 | -0.0268 ± 0.0003 | $7.51 \times 10^{-14}$ |
| 1025 | -0.0081 ± 0.0001 | -0.0094 ± 0.0002 | $9.31 \times 10^{-10}$ |
| 1079 | 0.0125 ± 0.0003 | 0.0105 ± 0.0003 | $7.15 \times 10^{-8}$ |
| 1374 | -0.0048 ± 0.0002 | -0.0071 ± 0.0002 | $1.87 \times 10^{-21}$ |
| 1604 | 0.0062 ± 0.0001 | 0.0081 ± 0.0002 | $3.64 \times 10^{-17}$ |
| 3027 | -0.0002 ± 0.0003 | 0.0090 ± 0.0005 | $1.58 \times 10^{-53}$ |
| 3110 | 0.0076 ± 0.0005 | 0.0191 ± 0.0006 | $3.22 \times 10^{-52}$ |

Abbreviations: AD, Alzheimer disease; FTD, frontotemporal dementia; SEM, standard error of the mean.
P values were calculated using 2-sided spectrum-level Welch independent-samples t tests. Because multiple Raman spectra were obtained from each participant, these comparisons represent exploratory spectrum-level analyses.

### eTable 3. Sex-Stratified Raman Intensities at Selected Wavenumbers in HC and Dementia Groups

#### A. Female Participants

| Wavenumber ($cm^{-1}$) | HC (mean ± SEM) | Dementia (mean ± SEM) | P value |
|---|---|---|---|
| 455 | 0.0190 ± 0.0003 | 0.0107 ± 0.0002 | $4.35 \times 10^{-88}$ |
| 532 | 0.0185 ± 0.0004 | 0.0330 ± 0.0005 | $5.80 \times 10^{-103}$ |
| 927 | 0.1534 ± 0.0010 | 0.1214 ± 0.0011 | $3.65 \times 10^{-89}$ |
| 973 | 0.0531 ± 0.0008 | 0.0771 ± 0.0009 | $5.04 \times 10^{-87}$ |
| 1255 | -0.0002 ± 0.0000 | -0.0013 ± 0.0001 | $1.45 \times 10^{-80}$ |
| 1567 | 0.0060 ± 0.0002 | 0.0088 ± 0.0002 | $7.74 \times 10^{-31}$ |
| 2863 | 0.0123 ± 0.0005 | 0.0040 ± 0.0003 | $2.54 \times 10^{-44}$ |
| 3027 | -0.0021 ± 0.0004 | 0.0033 ± 0.0003 | $2.31 \times 10^{-25}$ |

#### B. Male Participants

| Wavenumber ($cm^{-1}$) | HC (mean ± SEM) | Dementia (mean ± SEM) | P value |
|---|---|---|---|
| 655 | -0.0148 ± 0.0004 | -0.0201 ± 0.0003 | $1.37 \times 10^{-30}$ |
| 788 | -0.0210 ± 0.0004 | -0.0278 ± 0.0003 | $6.55 \times 10^{-36}$ |
| 1033 | -0.0043 ± 0.0004 | -0.0103 ± 0.0002 | $5.25 \times 10^{-32}$ |
| 1162 | -0.0191 ± 0.0003 | -0.0239 ± 0.0002 | $6.58 \times 10^{-39}$ |
| 1315 | -0.0095 ± 0.0002 | -0.0122 ± 0.0002 | $6.90 \times 10^{-19}$ |
| 1527 | 0.0001 ± 0.0002 | 0.0020 ± 0.0001 | $1.86 \times 10^{-18}$ |
| 2919 | 0.0196 ± 0.0007 | 0.0120 ± 0.0004 | $8.73 \times 10^{-19}$ |
| 2998 | 0.0023 ± 0.0005 | -0.0070 ± 0.0003 | $7.62 \times 10^{-51}$ |

Abbreviations: HC, cognitively healthy controls; SEM, standard error of the mean.
P values were calculated using 2-sided spectrum-level Welch independent-samples t tests. Because multiple Raman spectra were obtained from each participant, these comparisons represent exploratory spectrum-level analyses.

### eTable 4. Sex-Stratified Raman Intensities at Selected Wavenumbers in AD and FTD

#### A. Female Participants

| **Wavenumber ($cm^{-1}$)** | AD (mean ± SEM) | FTD (mean ± SEM) | P value |
|---|---|---|---|
| 468 | 0.0126 ± 0.0003 | 0.0097 ± 0.0003 | $3.11 \times 10^{-11}$ |
| 547 | 0.0436 ± 0.0005 | 0.0488 ± 0.0006 | $1.42 \times 10^{-11}$ |
| 904 | 0.0608 ± 0.0006 | 0.0647 ± 0.0007 | $1.71 \times 10^{-5}$ |
| 1147 | -0.0118 ± 0.0003 | -0.0137 ± 0.0003 | $1.23 \times 10^{-6}$ |
| 1293 | -0.0045 ± 0.0004 | -0.0093 ± 0.0004 | $3.44 \times 10^{-18}$ |
| 1361 | -0.0058 ± 0.0003 | -0.0094 ± 0.0002 | $3.06 \times 10^{-25}$ |
| 3025 | 0.0035 ± 0.0005 | 0.0101 ± 0.0005 | $1.81 \times 10^{-19}$ |
| 3110 | 0.0076 ± 0.0006 | 0.0200 ± 0.0006 | $2.15 \times 10^{-43}$ |

#### B. Male Participants

| **Wavenumber ($cm^{-1}$)** | **AD (mean ± SEM)** | **FTD (mean ± SEM)** | **P value** |
|---|---|---|---|
| 536 | 0.0304 ± 0.0006 | 0.0231 ± 0.0007 | $1.63 \times 10^{-15}$ |
| 775 | -0.0250 ± 0.0004 | -0.0305 ± 0.0005 | $8.18 \times 10^{-17}$ |
| 885 | 0.0465 ± 0.0005 | 0.0374 ± 0.0007 | $8.92 \times 10^{-23}$ |
| 1033 | -0.0090 ± 0.0003 | -0.0122 ± 0.0003 | $2.13 \times 10^{-15}$ |
| 1473 | -0.0002 ± 0.0002 | -0.0025 ± 0.0003 | $1.47 \times 10^{-14}$ |
| 1604 | 0.0055 ± 0.0002 | 0.0083 ± 0.0003 | $6.70 \times 10^{-16}$ |
| 2998 | -0.0034 ± 0.0004 | -0.0125 ± 0.0005 | $3.33 \times 10^{-44}$ |
| 3029 | -0.0006 ± 0.0005 | 0.0124 ± 0.0008 | $8.00 \times 10^{-39}$ |

Abbreviations: AD, Alzheimer disease; FTD, frontotemporal dementia; SEM, standard error of the mean.
P values were calculated using 2-sided spectrum-level Welch independent-samples t tests. Because multiple Raman spectra were obtained from each participant, these comparisons represent exploratory spectrum-level analyses.

### eFigure 1. Raman Spectral Preprocessing Workflow for Sweat EV Samples

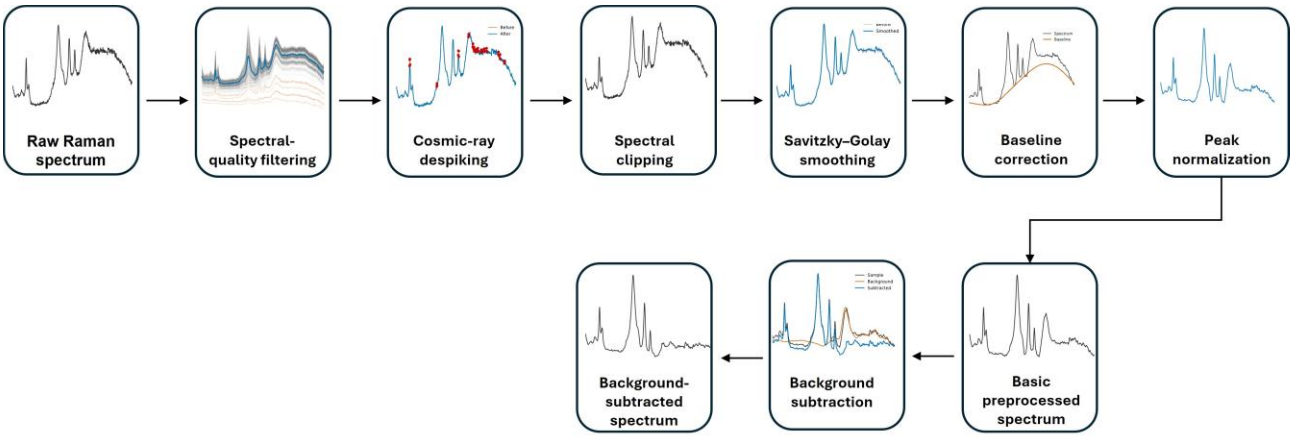


Schematic overview of Raman spectral preprocessing applied to sweat extracellular vesicle (EV) spectra. Raw Raman spectra acquired from selected sample regions underwent spectral quality filtering, cosmic-ray despiking, spectral clipping, Savitzky-Golay smoothing, baseline correction, and peak-based normalization to generate preprocessed spectra. The mean background spectrum was subsequently subtracted to generate the background-subtracted spectra used for downstream statistical and machine-learning analyses.
Abbreviation: EV, extracellular vesicle.

### eFigure 2. Characterization of Sweat Extracellular Vesicles by Nanoparticle Tracking Analysis and Transmission Electron Microscopy

#### A = NTA size-distribution profiles

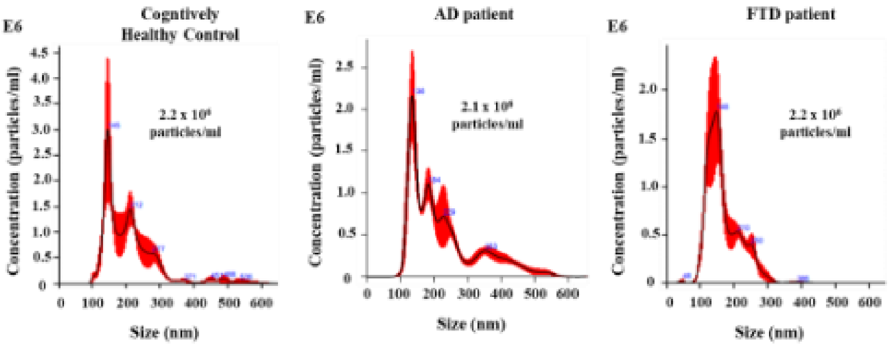


#### B = TEM images

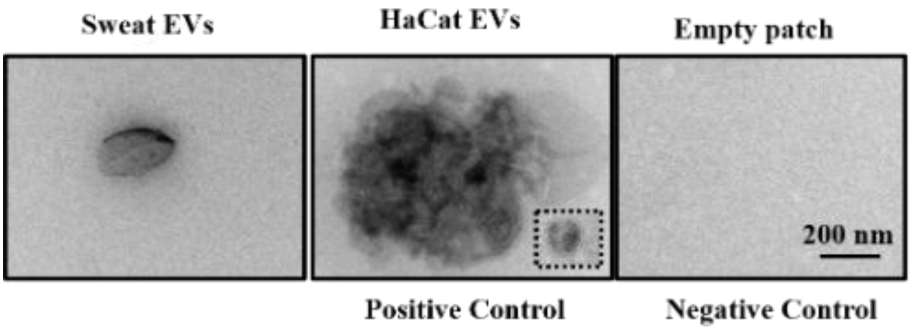


**A,** Representative nanoparticle tracking analysis (NTA) size-distribution profiles of sweat extracellular vesicles (EVs) from cognitively healthy controls (HC) and participants with Alzheimer disease (AD) and frontotemporal dementia (FTD). **B,** Representative transmission electron microscopy (TEM) images of negatively stained EVs isolated from sweat-patch samples and from the human keratinocyte cell line HaCaT, which served as a positive control. Material processed from an unused sweat patch that underwent the same isolation and processing procedures served as the negative control. The dashed rectangle in the HaCaT image highlights an individual EV. Scale bar, 200 nm.

Abbreviations: AD, Alzheimer disease; EV, extracellular vesicle; FTD, frontotemporal dementia; HC, cognitively healthy controls; NTA, nanoparticle tracking analysis; TEM, transmission electron microscopy.

### eFigure 3. Raman Intensity Distributions at Selected Wavenumbers in Cognitively Healthy Controls and Participants With Dementia

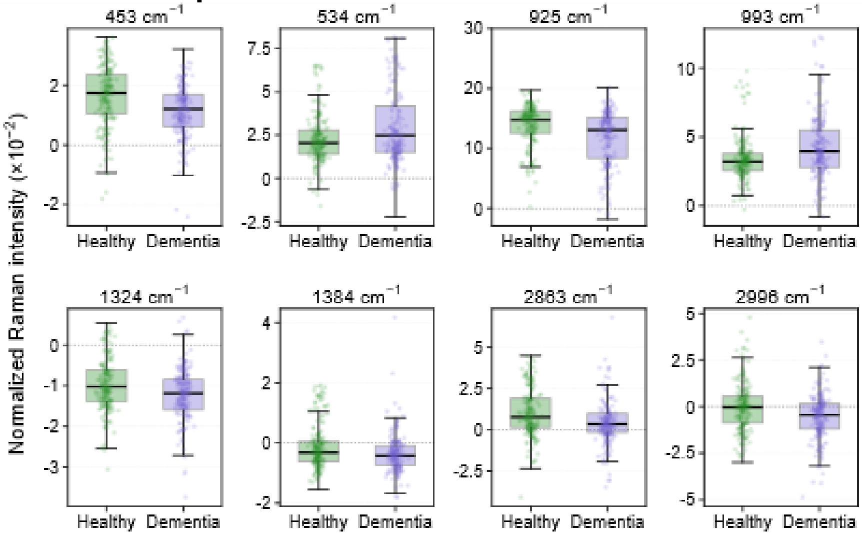


Boxplots show normalized Raman intensities at the 8 selected wavenumbers identified in the HC-vs-dementia analysis in Figure 1C: 453, 534, 925, 993, 1324, 1384, 2863, and 2996 cm$^{-1}$. Boxes indicate the interquartile range, central horizontal lines indicate the median, and jittered points represent a randomly selected subset of spectra for visualization; all available spectra were included in the boxplot calculations. Raman intensities are displayed as ×10$^{-2}$, with the y-axis range adjusted separately for each wavenumber for visualization. Corresponding mean (SEM) values and exact *P* values from spectrum-level 2-sided Welch independent-samples *t* tests are provided in eTable 1.

Abbreviations: HC, cognitively healthy controls; SEM, standard error of the mean.

### eFigure 4. Raman Intensity Distributions at Selected Wavenumbers in AD and FTD

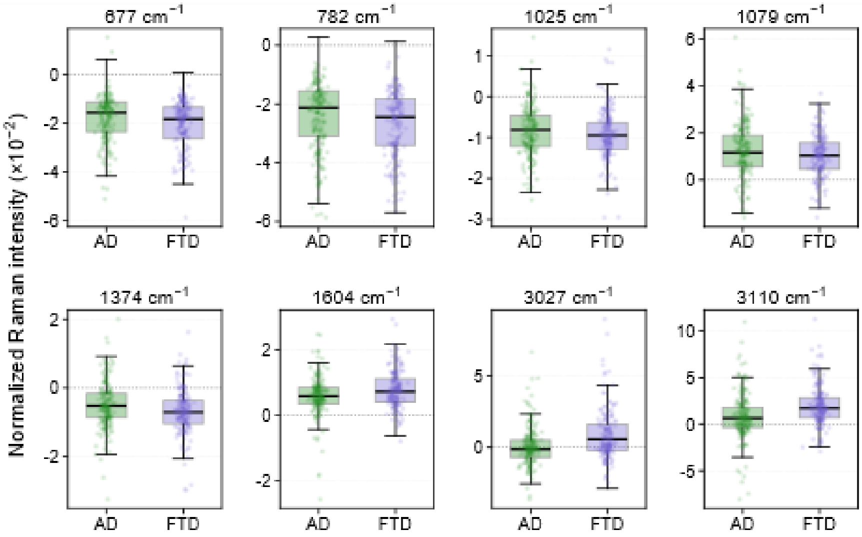


Boxplots show normalized Raman intensities at the 8 selected wavenumbers identified in the AD-vs-FTD analysis in Figure 1D: 677, 782, 1025, 1079, 1374, 1604, 3027, and 3110 cm$^{-1}$. Boxes indicate the interquartile range, central horizontal lines indicate the median, and jittered points represent a randomly selected subset of spectra for visualization; all available spectra were included in the boxplot calculations. Raman intensities are displayed as ×10$^{-2}$, with the y-axis range adjusted separately for each wavenumber for visualization. Corresponding mean (SEM) values and exact *P* values from spectrum-level 2-sided Welch independent-samples *t* tests are provided in eTable 2.

Abbreviations: AD, Alzheimer disease; FTD, frontotemporal dementia; SEM, standard error of the mean.

### eFigure 5. Sex-Stratified Raman Intensity Distributions in Cognitively Healthy Controls and Participants With Dementia

#### A, Female participants

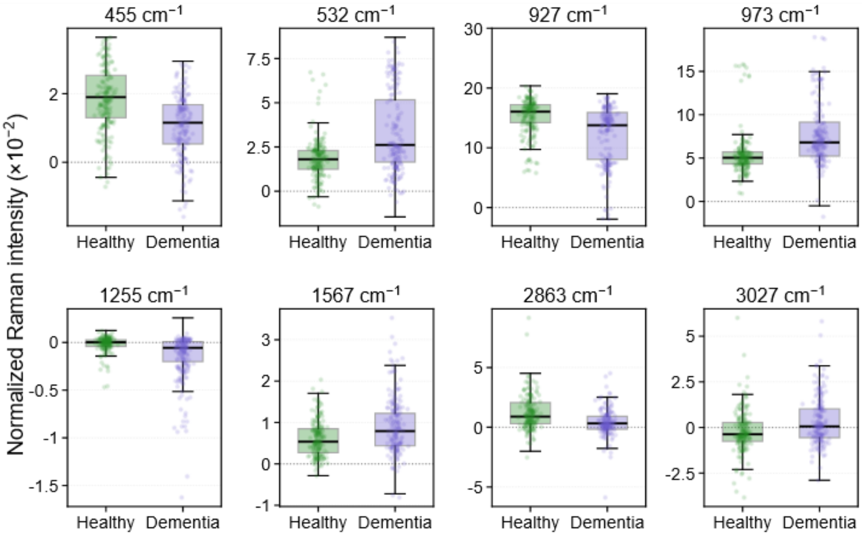


#### B, Male participants

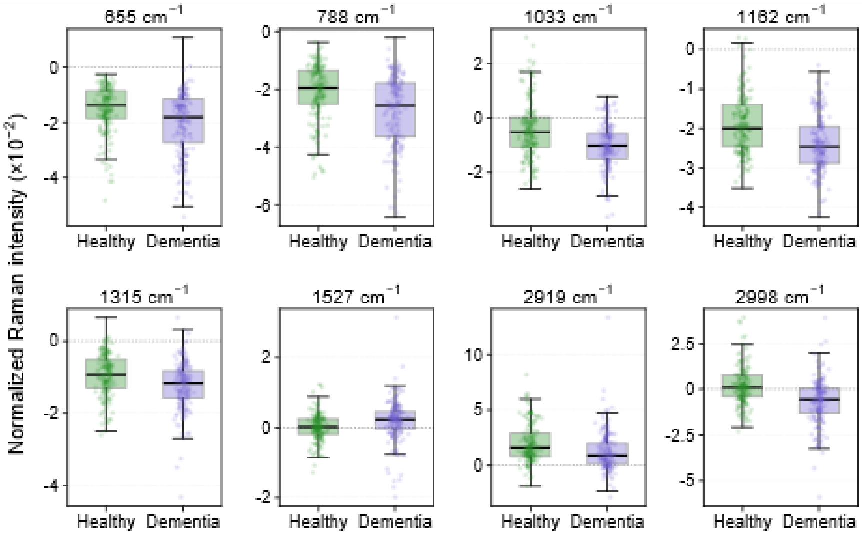


**A,** Normalized Raman intensity distributions in female participants at the 8 selected wavenumbers identified in Figure 2A: 455, 532, 927, 973, 1255, 1567, 2863, and 3027 $cm^{-1}$. **B,** Corresponding distributions in male participants at the 8 selected wavenumbers identified in Figure 2B: 655, 788, 1033, 1162, 1315, 1527, 2919, and 2998 $cm^{-1}$. Boxes indicate the interquartile range, central horizontal lines indicate the median, and jittered points represent a randomly selected subset of spectra for visualization; all available spectra were included in the boxplot calculations. Raman intensities are displayed as $\times10^{-2}$, with the y-axis range adjusted separately for each wavenumber for visualization. Corresponding mean (SEM) values and exact *P* values from spectrum-level 2-sided Welch independent-samples *t* tests are provided in eTable 3.

Abbreviations: HC, cognitively healthy controls; SEM, standard error of the mean.

### eFigure 6. Sex-Stratified Raman Intensity Distributions in AD and FTD

#### A, Female participants

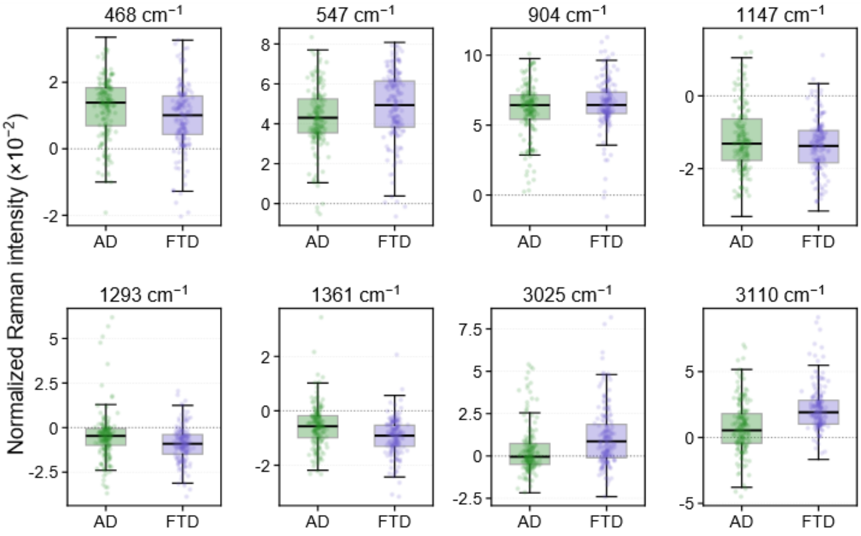


#### B, Male participants

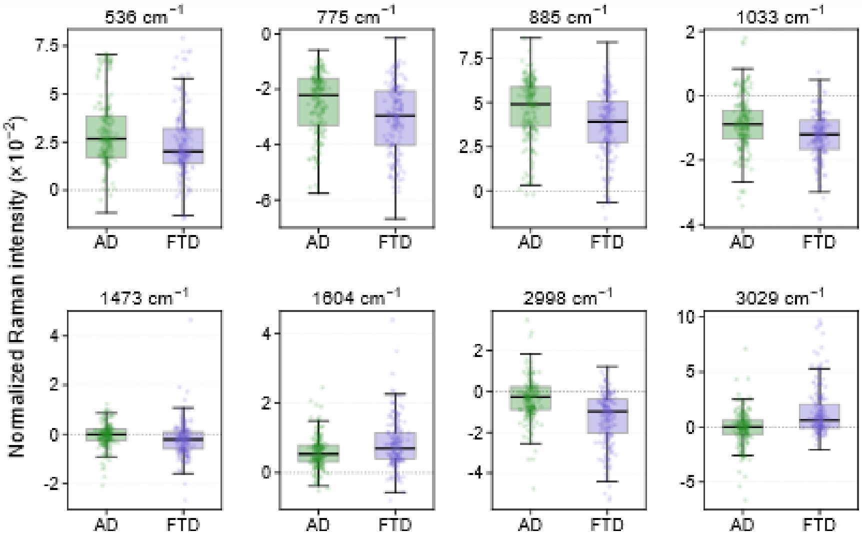


**A,** Normalized Raman intensity distributions in female participants at the 8 selected wavenumbers identified in Figure 2C: 468, 547, 904, 1147, 1293, 1361, 3025, and 3110 $cm^{-1}$. **B,** Corresponding distributions in male participants at the 8 selected wavenumbers identified in Figure 2D: 536, 775, 885, 1033, 1473, 1604, 2998, and 3029 $cm^{-1}$. Boxes indicate the interquartile range, central horizontal lines indicate the median, and jittered points represent a randomly selected subset of spectra for visualization; all available spectra were included in the boxplot calculations. Raman intensities are displayed as $\times10^{-2}$, with the y-axis range adjusted separately for each wavenumber for visualization. Corresponding mean (SEM) values and exact *P* values from spectrum-level 2-sided Welch independent-samples *t* tests are provided in eTable 4.
Abbreviations: AD, Alzheimer disease; FTD, frontotemporal dementia; SEM, standard error of the mean.

### eFigure 7. Sex-Stratified HC vs Dementia Classification in Full and Balanced Cohorts

**A, Full cohort**

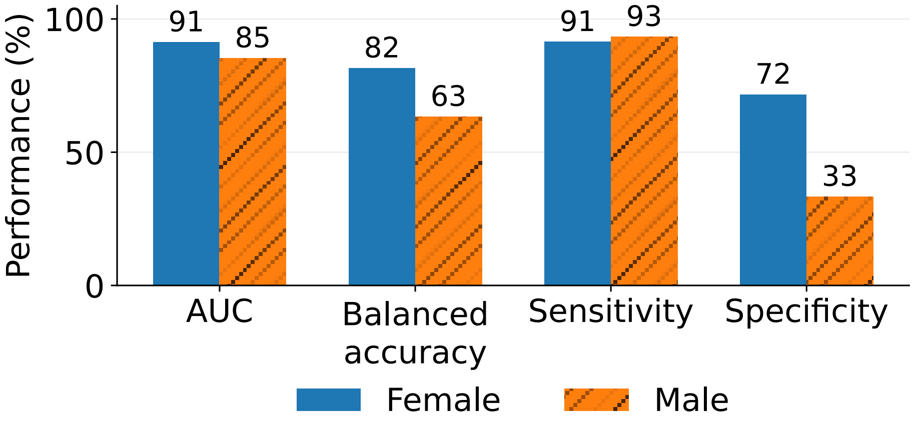


**B, Balanced cohort**

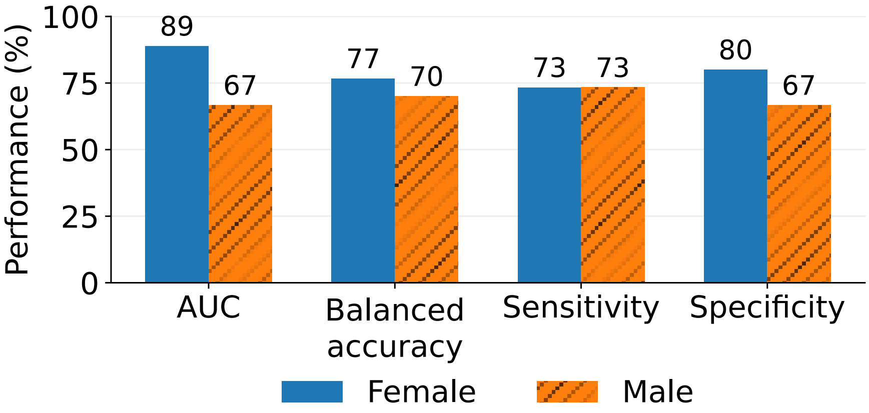


**A,** Classification performance for female and male participants in the full HC-vs-dementia cohort. **B,** Classification performance for female and male participants in the matched balanced HC-vs-dementia cohort. Models used background-subtracted sweat extracellular vesicle (EV) Raman spectra and stratified group cross-validation with participants as the grouping variable. Bars show performance for the top-ranked model configuration. AUC values are displayed as percentages to permit comparison with balanced accuracy, sensitivity, and specificity. Sensitivity represents correct identification of participants with dementia, and specificity represents correct identification of HC participants. Values above bars indicate rounded percentages.
Abbreviations: AUC, area under the receiver operating characteristic curve; EV, extracellular vesicle; HC, cognitively healthy controls.

**eFigure 8. AD vs FTD Classification Overall and by Sex**

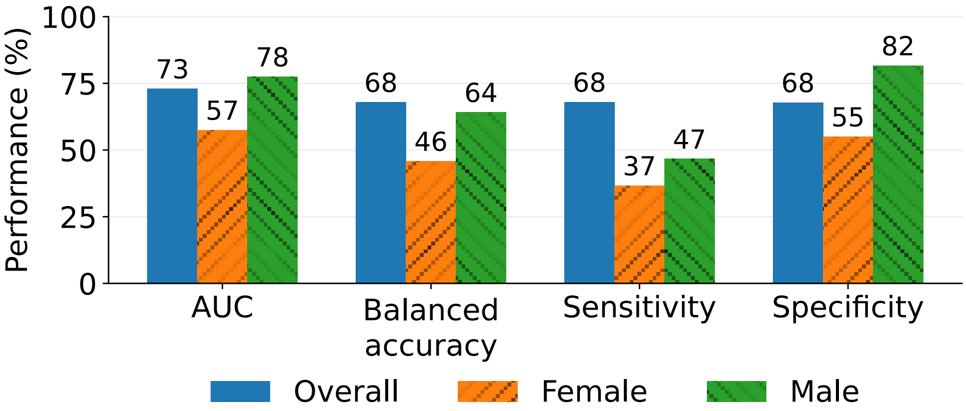


Classification performance for the overall, female, and male AD-vs-FTD analyses using background-subtracted sweat extracellular vesicle (EV) Raman spectra and stratified group cross-validation with participants as the grouping variable. Bars show performance for the top-ranked model configuration. AUC values are displayed as percentages to permit comparison with balanced accuracy, sensitivity, and specificity. Sensitivity represents correct identification of FTD, and specificity represents correct identification of AD. Values above bars indicate rounded percentages.

Abbreviations: AD, Alzheimer disease; AUC, area under the receiver operating characteristic curve; EV, extracellular vesicle; FTD, frontotemporal dementia.
